# Establishing wastewater metagenomics as a quantitative pathogen monitoring tool with normalization

**DOI:** 10.64898/2026.07.14.26356442

**Authors:** Lennart Justen, Alessandro Zulli, Rose S. Kantor, Rebecca Y. Linfield, Leon S. Moskatel, Daniel Cunningham-Bryant, Jeff Kaufman, Marc C. Johnson, Michael R. McLaren, Pardis C. Sabeti

## Abstract

Wastewater metagenomic sequencing (WW-MGS) enables simultaneous detection of hundreds of pathogens, but its use for quantitative pathogen tracking has not been robustly validated. Like wastewater PCR (WW-PCR), WW-MGS is affected by biases from variable fecal dilution and sample processing, but must additionally contend with the compositional structure of sequencing data, where a taxon’s apparent abundance depends on the abundance of every other taxon in the sample. Simple summaries such as a pathogen’s fraction of total reads may therefore be poorly suited to quantitative use. We retrospectively evaluated seven normalization approaches that attempt to control for these sources of bias against a baseline of total read relative abundance, using 1,425 samples from the CASPER consortium spanning 25 U.S. sites. Each approach was compared against WW-PCR and clinical data across eight total pathogens. Among the normalization strategies we evaluated, tobamovirus markers, diet-derived plant viruses abundant in human stool, performed best. Normalizing WW-MGS data by tobamovirus-genus counts improved median site concordance for 18 of 19 pathogen and comparison-source combinations. Gains were largest for year-round-circulating SARS-CoV-2 and norovirus and smaller for sharply seasonal pathogens such as influenza and respiratory syncytial virus, where baseline concordance was already high. Tobamovirus normalization rarely degraded concordance, with median gains roughly five times larger than median losses. Tobamovirus-normalized WW-MGS reached clinical concordance comparable to targeted WW-PCR, supporting its use as a quantitative trend-monitoring tool alongside pathogen-agnostic detection.

## Introduction

Wastewater monitoring provides cost-effective, population-level pathogen tracking that is independent of healthcare-seeking behavior and clinical testing patterns (Diamond et al., 2022; Peccia et al., 2020). These advantages drove widespread use of wastewater PCR (WW-PCR) in the United States during the COVID-19 pandemic to track SARS-CoV-2 levels, especially as in-person testing rates fell, and national programs such as the CDC National Wastewater Surveillance System (NWSS) and WastewaterSCAN have since extended WW-PCR monitoring to dozens of additional pathogens (Adams et al., 2024; Boehm et al., 2024). While the temporal pathogen abundance trends measured by WW-PCR correlate well with clinical surveillance and are generally considered reliable proxies of infection rates, WW-PCR signals remain sensitive to bias from fluctuations in pathogen nucleic acid concentrations unrelated to human shedding dynamics. These include sample dilution from stormwater or other sewer inflow and variation in sample processing and nucleic acid recovery.

To reduce variation in measured pathogen concentrations due to outside effects, researchers often normalize WW-PCR signals (Ahmed et al., 2026). The most common approach uses a human-associated fecal marker that is co-shed and co-processed with the target pathogen, under the assumption that the marker itself is shed at a relatively stable per-capita rate. The most widely used marker is pepper mild mottle virus (PMMoV), a plant virus abundant in human stool through diet. In this approach, the raw pathogen WW-PCR concentration (copies/L or copies/g) is divided by the PMMoV concentration in the same sample and reported as pathogen copies per PMMoV copy (Ahmed et al., 2026; Symonds et al., 2018; Wolfe et al., 2021). However, evidence that PMMoV normalization improves concordance with clinical data is mixed and largely limited to analysis of SARS-CoV-2. PMMoV normalization is generally beneficial in primary settled solids (Graham et al., 2021; Wolfe et al., 2021), but in influent samples it often has no effect or reduces correlations with clinical data, with flow-rate and chemical measures of fecal strength (e.g., ammonia, total nitrogen, caffeine) sometimes performing better (Dhiyebi et al., 2023; Duvallet et al., 2022; Feng et al., 2021; Greenwald et al., 2021; Hoar et al., 2022; Hsu et al., 2022; Maal-Bared et al., 2023; Schill et al., 2023). Alternative fecal markers including tomato brown rugose fruit virus (ToBRFV), which exhibits higher abundance and potentially lower temporal variability than PMMoV in wastewater (Natarajan et al., 2023), show promise but have not been systematically evaluated as normalization targets for pathogen trend data.

More recently, wastewater metagenomic sequencing (WW-MGS) has demonstrated potential as a pathogen-agnostic surveillance tool, enabling simultaneous detection of hundreds of viral taxa without prior assay design or targeting (Justen et al., 2026; Rushford et al., 2025; Worp et al., 2025). Unlike WW-PCR, which measures absolute nucleic acid concentrations, sequencing returns a finite number of reads per sample and is therefore compositional: a given taxon’s read count depends not only on how much of that taxon was present, but also on the total number of reads generated and on the abundance of every other taxon sequenced alongside it (Gloor et al., 2017; McLaren et al., 2019). Reporting relative abundance, the fraction of reads assigned to a taxon, controls for variation in sequencing depth but not for changes in the rest of the library. If unrelated taxa become more abundant, the focal taxon’s relative abundance can fall even when its absolute abundance is unchanged. In wastewater RNA sequencing, for instance, bacterial ribosomal RNA (rRNA) dominates and often makes up more than half of all reads, so variation in rRNA content driven by sample processing, reagent performance, or environmental microbial dynamics can substantially alter apparent viral relative abundance. Compositionality thus introduces a second, arguably more severe layer of distortion on top of the biases already affecting WW-PCR, but one that can in principle be addressed by normalization to a marker taxon also present in the WW-MGS data (Gloor et al., 2017).

Here, we use data from the Coalition for Agnostic Sequencing of Pathogens from Environmental Reservoirs (CASPER), a U.S.-based WW-MGS network that has generated the largest publicly available untargeted WW-MGS dataset to date (Justen et al., 2026), to evaluate how different normalization strategies affect concordance between WW-MGS pathogen abundance signals and clinical and WW-PCR comparators. We assess seven normalization approaches across eight pathogens and 25 sites, taking advantage of the fact that sequencing measures candidate normalization markers alongside pathogen targets at no additional cost. The strategies span single-target fecal markers (PMMoV, ToBRFV), broader read denominators (non-rRNA reads), and composite or aggregate indicators (tobamovirus genus, *Crassvirales*, human reads, and a strict anaerobic gut bacteria composite, SABC). We compare each approach against three established epidemiological signals: WW-PCR from NWSS and WastewaterSCAN, clinical encounters from Epic Cosmos, and hospital respiratory admissions from the National Healthcare Safety Network (NHSN), and characterize the impact of normalization. The findings apply most directly to WW-MGS but also provide insight into normalization strategies for WW-PCR monitoring. This, to our knowledge, represents the first systematic evaluation of normalization for WW-MGS pathogen quantification.

## Methods

### Data sources

#### Wastewater metagenomic sequencing (WW-MGS)

We analyzed publicly available WW-MGS data generated by the CASPER network (NCBI BioProject accession PRJNA1247874) between December 2023 and March 2026; sample collection, processing, and high-level taxonomic composition are described in detail in Justen et al. (2026) (Table 1). Briefly, CASPER partners collected 24-hour composite influent samples at approximately weekly intervals from municipal wastewater treatment plants across the U.S and shipped them to the University of Missouri (MU) processing laboratory for viral concentration, nucleic acid extraction, cDNA synthesis, and Illumina library preparation. The resulting libraries underwent deep, untargeted RNA sequencing on Illumina NovaSeq instruments (2×150 bp paired-end reads), targeting approximately one billion read pairs per sample. Although the protocol targets RNA, it also captures some DNA and DNA viruses.

**Table 1.** CASPER sampling sites included in the wastewater metagenomic sequencing (WW-MGS) normalization analysis. The table lists site location, estimated population served, sample type, collection start date, and number of samples in this data release. Population estimates are approximate and based on sewershed coverage data from the National Wastewater Surveillance System (NWSS) where available; “–” indicates sites that opted to keep population estimates anonymous. WWTP, wastewater treatment plant.

| Sampling site | Location type | Sample type | Population served | Collection start date | Samples |
| --- | --- | --- | --- | --- | --- |
| **Northeast** |  |  |  |  |  |
| Boston Deer Island Treatment Plant (DITP) North, MA | WWTP | 24h composite | 1.1M | 2024-03-20 | 76 |
| Boston DITP South, MA | WWTP | 24h composite | 1.4M | 2024-03-20 | 76 |
| **Midwest** |  |  |  |  |  |
| Chicago (CHI-A), IL | WWTP | 24h composite | 1.3M | 2024-09-08 | 82 |
| Chicago (CHI-B), IL | WWTP | 24h composite | 1.1M | 2024-09-08 | 76 |
| Chicago (CHI-D1), IL | WWTP | 24h composite | 1.1M | 2024-09-15 | 76 |
| Chicago (CHI-D2), IL | WWTP | 24h composite | 1.1M | 2024-09-15 | 76 |
| Ottumwa Water Pollution Control Facility (WPCF), IA | WWTP | 24h composite | 26k | 2025-02-19 | 58 |
| Bissell Point WWTP, MO | WWTP | 24h composite | 310k | 2025-06-03 | 46 |
| Coldwater Creek WWTP, MO | WWTP | 24h composite | 150k | 2025-06-03 | 48 |
| Columbia WWTP, MO | WWTP | 24h composite | 120k | 2023-12-26 | 119 |
| Kansas City Blue River WWTP, MO | WWTP | 24h composite | 490k | 2025-08-12 | 34 |
| Kansas City Westside WWTP, MO | WWTP | 24h composite | 61k | 2025-10-01 | 25 |
| Lemay WWTP, MO | WWTP | 24h composite | 450k | 2025-06-03 | 46 |
| Milan Wastewater Treatment Facility (WWTF), MO | WWTP | 24h composite | 1.8k | 2025-02-24 | 39 |
| Missouri River WWTP, MO | WWTP | 24h composite | 180k | 2025-06-20 | 42 |
| Monett WWTP, MO | WWTP | 24h composite | 9.1k | 2025-06-10 | 42 |
| **South** |  |  |  |  |  |
| Miami-Dade Central District Wastewater Treatment Plant (CDWWTP), FL | WWTP | 24h composite | 900k | 2026-01-12 | 12 |
| Central Oklahoma (OK-A), OK | WWTP | 24h composite | 110k | 2025-08-18 | 32 |
| Central Oklahoma (OK-B), OK | WWTP | 24h composite | 440k | 2025-08-18 | 32 |
| **West** |  |  |  |  |  |
| Ontario, CA | WWTP | 24h composite | 890k | 2025-08-25 | 40 |
| Palo Alto Regional Water Quality Control Plant (RWQCP), CA | WWTP | 24h composite | 240k | 2025-08-25 | 31 |
| Riverside Water Quality Control Plant (WQCP), CA | WWTP | 24h composite | 350k | 2024-12-09 | 121 |
| Sacramento, CA | WWTP | 24h composite | 1.6M | 2025-08-28 | 30 |
| Southern California, CA | WWTP | 24h composite | – | 2025-04-16 | 96 |
| West Boise and Lander Street Water Renewal Facility (Boise RWF), ID | WWTP | 24h composite | 270k | 2024-11-04 | 70 |

For this analysis, we restricted the dataset to MU-processed CASPER sites with 24-hour composite municipal wastewater treatment plant (WWTP) samples that have matching WW-PCR or clinical comparison data for ten or more overlapping Morbidity and Mortality Weekly Report (MMWR) weeks (Table 1) for at least one of the eight analyzed pathogens. After applying these criteria, the analysis includes 1,425 samples from 25 unique catchments across eight U.S. states. Sequencing depth across samples had a median of 0.95 billion read pairs (range 0.05B–2.63B; Supplementary Figure S1), corresponding to approximately 290 gigabases of data per sample.

We analyzed sequencing data using SecureBio’s Viral Metagenomics Pipeline (v3.0.1; index version 20250825; github.com/securebio/nao-mgs-workflow; (Bradshaw et al., 2026)). The pipeline performs two taxonomic profiling workflows, both described in Justen et al. (2026). First, the pipeline performed shallow taxonomic profiling on a random subsample of read pairs from each sequencing lane, yielding a median of 8.0 million subsampled read pairs (range 2.0M–32.0M). The workflow quality-filtered subsampled reads, partitioned them into ribosomal and non-ribosomal fractions using BBDuk against the SILVA rRNA databases (Chuvochina et al., 2025), and classified them using Kraken2 (Wood et al., 2019) with the Standard reference database (k2_standard_20250714). This profiling provided the rRNA fraction and abundance estimates for all normalization markers. Second, the pipeline processed all reads through a dedicated vertebrate-infecting virus workflow that uses k-mer screening, alignment to a curated vertebrate-infecting virus genome database with Bowtie2 (Langmead & Salzberg, 2012), contaminant filtering, and taxonomic assignment via a lowest common ancestor algorithm to sensitively identify pathogen reads. Viral host annotation uses a custom algorithm that propagates host information through the NCBI viral taxonomy based on Virus-Host DB annotations (Mihara et al., 2016), as described in Justen et al. (2026).

#### Wastewater PCR (WW-PCR)

We obtained WW-PCR measurements from WastewaterSCAN and NWSS. For sites with ten or more weeks of overlapping WW-MGS and WastewaterSCAN coverage (5 of 25 sites; Supplementary Table S1), we used droplet digital RT-PCR measurements from a published WastewaterSCAN data release (Boehm et al., 2026), which provides concentrations in gene copies per gram dry weight of wastewater solids for all eight pathogens analyzed: SARS-CoV-2, influenza A, influenza B, respiratory syncytial virus (RSV), human metapneumovirus (HMPV), norovirus GII, rotavirus, and enterovirus D68 (EV-D68). WastewaterSCAN data are shared under a CC BY-NC 4.0 license and were collected as part of a partnership between Stanford University, Emory University, and Verily.

For sites without WastewaterSCAN coverage but with sufficient state- or territory-sourced NWSS data overlapping the CASPER sampling period (11 of 25 sites), we obtained SARS-CoV-2, influenza A, and RSV PCR concentrations, reported as copies per liter, from the CDC NWSS data portal (Adams et al., 2024). We downloaded NWSS data via the CDC Socrata API from public datasets for SARS-CoV-2 (dataset j9g8-acpt; (CDC, 2025c)), influenza A (dataset ymmh-divb; (CDC, 2025a)), and RSV (dataset 45cq-cw4i; (CDC, 2025b)). Local partners helped match sewershed identifiers from both sources to CASPER sampling sites (Supplementary Table S1). Nine sites without usable WW-PCR comparison data were excluded from WW-PCR analyses but retained for clinical-comparator analyses when sufficient overlapping clinical data were available.

For evaluating WW-MGS normalization approaches, we use raw unnormalized WW-PCR concentrations, reported as gene copies per dry weight or copies per liquid volume, as the primary comparison target. Although NWSS also reports PMMoV-normalized and flow-normalized concentrations, and WastewaterSCAN provides PMMoV measurements for normalization, using the raw WW-PCR signal avoids confounding the evaluation of WW-MGS normalization with WW-PCR-side normalization choices. In our data, PMMoV- and flow-normalized WW-PCR concentrations correlated with clinical comparators similarly to raw WW-PCR concentrations and had little effect on the conclusions (Supplementary Figure S2), consistent with previous work (Chan et al., 2025).

#### Clinical

##### Epic Cosmos (Cosmos)

We obtained clinical encounter data from Cosmos (Epic Systems Corporation, Verona, Wisconsin), a dataset created in collaboration with a community of health systems using Epic. As of April 2026, Cosmos represented more than 301 million patient records from over 2,050 hospitals and 47,100 clinics from all 50 states, D.C., Canada, Lebanon, and Saudi Arabia. We obtained weekly state-level encounter counts from Cosmos for the eight states containing CASPER sites, covering November 1, 2023 through March 30, 2026. We exported data on April 12, 2026.

For SARS-CoV-2, RSV, HMPV, norovirus, and rotavirus, we used encounters associated with pathogen-specific International Classification of Diseases, 10th edition (ICD-10) diagnostic codes. For influenza A and influenza B, we used encounters with positive laboratory test results, because ICD-10 codes for influenza do not distinguish between types. We also compared a combined influenza A and B WW-MGS signal against encounter counts from the ICD-10-based J09–J11 codes, which captures both types without stratification. Supplementary Table S2 provides ICD-10 code mappings, laboratory test specifications, and pathogen-by-pathogen taxonomic matching between WW-MGS, PCR, and clinical assay specificity.

Cosmos suppresses cell values representing ten or fewer encounters to protect patient privacy. We treated suppressed values as zero, preserving low-incidence weeks as informative observations in the correlation analysis rather than excluding them. We confirmed empirically that this choice did not drive our conclusions compared to excluding suppressed values as missing values (*NaN*) (Supplementary Figure S3). Across the eight pathogen × state combinations entering the Cosmos analyses, 1,475 of 4,176 weekly observations (35.3%) were suppressed; suppression was rare for SARS-CoV-2 (1.0%) and combined influenza (0.6%) but high for HMPV (62.8%), norovirus (50.4%), and rotavirus (91.4%) (Supplementary Figure S4). The Stanford University Institutional Review Board determined that this project does not meet the definition of human subject research (Protocol 87814). Cosmos data are deidentified prior to aggregation and no individual-level records were accessed.

##### National Healthcare Safety Network (NHSN) Hospital Respiratory Data (HRD)

The CDC’s NHSN collects weekly counts of new hospital admissions of patients with laboratory-confirmed COVID-19, influenza, and RSV. NHSN counts an admission when an inpatient has a positive laboratory test for the pathogen within the 14 days preceding admission, whether the test was performed in an inpatient or outpatient setting; admissions without a positive test, and positive tests not associated with an inpatient admission, are not counted (CDC, 2026). We use publicly available NHSN HRD data aggregated to the state level (dataset ua7e-t2fy; (CDC, 2024)). NHSN reports laboratory-confirmed influenza as a single count combining influenza A and B, so we compared NHSN influenza admissions against a combined-influenza WW-MGS signal (Supplementary Table S2). Hospital reporting of HRD became mandatory for acute-care and critical-access hospitals on November 1, 2024; data from the earlier voluntary reporting period are less complete.

#### Normalization methods

We evaluated seven candidate normalization approaches for WW-MGS pathogen abundance data, comparing each against a baseline of total read relative abundance. Each approach divides pathogen read counts by a different denominator intended to control for variation unrelated to pathogen shedding dynamics. We define read count as the number of read pairs assigned to a taxon, without deduplication. We selected normalization strategies based on prior literature, established WW-PCR normalization practices, and biological reasoning about which marker types might stably reflect human fecal input, including gut-associated, diet-derived, and host derived signals.

For a given sample, let *N*_*p*_ denote the number of reads assigned to pathogen *p* by the vertebrate-infecting virus pipeline, and let *N*_*total*_ denote the total number of read pairs in the sequencing library. Shallow taxonomic profiling estimates the rRNA fraction *f*_*rRNA*_ and normalization marker counts from a random subsample of *N*_*profiled*_ read pairs per sample. Because profiling uses a subsample, we scaled marker counts to library depth before using them as denominators. Specifically, we multiplied raw marker counts from profiling by *N*_*total*_/*N*_*profiled*_ to obtain estimated library-wide counts. We calculated the estimated number of non-ribosomal read pairs as *N*_*non−rRNA*_ = *N*_*total*_ (1 − *f*_*rRNA*_). In the formulas below, all marker counts refer to these scaled, library-wide estimates. A sample with a zero or undefined marker denominator produced *NaN* and was excluded from that normalization’s analysis; zero denominator drop counts are reported in Supplementary Table S3 and never exceeded 5 out of 1,425 samples across all denominator choices.

##### Total read relative abundance (baseline)

Pathogen reads divided by total reads:

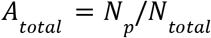

This approach applies no normalization beyond accounting for sequencing depth and serves as the baseline against which we compared all other approaches.

##### Non-rRNA relative abundance

Pathogen reads divided by estimated non-ribosomal reads:

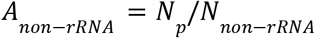

Ribosomal RNA, predominantly bacterial in origin, constitutes the majority of reads in wastewater metatranscriptomic libraries and varies substantially across samples and sites (Justen et al., 2026). Fluctuations in rRNA fraction driven by differences in ribodepletion efficiency, environmental microbial dynamics, or sample composition are a primary source of compositional variation affecting all non-rRNA taxa. Non-rRNA normalization provides a first-order compositional correction that does not depend on any specific marker organism.

##### Fecal indicator viruses: pepper mild mottle virus (PMMoV), tomato brown rugose fruit virus (ToBRFV), and tobamovirus genus

Pathogen reads divided by fecal indicator virus reads:

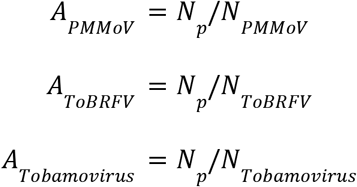

PMMoV and ToBRFV are diet-associated tobamoviruses shed in high concentrations in human feces after consumption of pepper- and tomato-containing foods, respectively, (Colson et al., 2010; Hamza et al., 2011; Persson et al., 2026; Zhang et al., 2006), and they are exceptionally stable in the environment (Bačnik et al., 2020; Mehle et al., 2023). Rosario et al. (2009) first proposed PMMoV as a fecal pollution indicator and PMMoV is now widely used in wastewater surveillance (Ahmed et al., 2026; Pappu et al., 2026; Symonds et al., 2018); CDC NWSS uses PMMoV normalization for WW-PCR data at many sites. ToBRFV has been proposed as an alternative fecal indicator because it can be more abundant and less temporally variable than PMMoV in wastewater (Natarajan et al., 2023; Rushford et al., 2025). The virus has also spread rapidly through commercial tomato production globally since its first reports in 2014–2015 (Luria et al., 2017; Salem et al., 2016).

Tobamovirus genus normalization uses all reads classified to the genus *Tobamovirus*, which encompasses PMMoV, ToBRFV, and other diet-associated plant viruses, including cucumber green mottle mosaic virus, tobacco mild green mosaic virus, and tomato mosaic virus (Rothman & Whiteson, 2022). By aggregating across related diet-associated markers, this approach may provide a more stable denominator than relying on any single tobamovirus species.

##### Human-derived reads

Pathogen reads divided by non-ribosomal human reads:

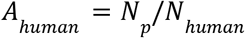

Human reads provide a direct marker of human biological input to the wastewater stream through fecal matter, as well as other sewer-bound sources such as skin cells, urine, and respiratory secretions. However, in the Kraken2 Standard database used for shallow taxonomic profiling, humans are the only eukaryote. As a result, some non-human animal reads may be assigned to the human clade and inflate human clade counts.

##### Human gut microbe markers: *Crassvirales* and strict anaerobe bacterial composite (SABC)

Non-rRNA normalization provides a broad compositional correction, but it does not specifically measure human fecal input. We therefore also evaluated endogenous human gut microorganisms as more specific fecal normalization markers. *Crassvirales* and SABC represent gut-associated microbial signals that may better track human fecal contribution than broad bacterial or non-rRNA read fractions, though they are less abundant than diet-derived tobamovirus markers..

*Crassvirales*. Bacteriophages of the order *Crassvirales*, including the prototypical crAssphage, are among the most abundant components of the mammalian gut DNA virome and infect bacteria of the phylum Bacteroidota, which dominate the human gut microbiome (Guerin et al., 2018; Stachler et al., 2017). Although the order as a whole is found across mammalian guts and other environments, the prototypical crAssphage is highly specific to human feces and has been used as a human fecal pollution marker (Stachler et al., 2017). We used the order-level *Crassvirales* marker because counts were approximately 14-fold more abundant than crAssphage counts (210 median read pairs per shallow taxonomic profiling subsample versus 15), yielding more stable denominators at the cost of some human specificity.

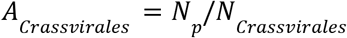

*SABC*. The SABC sums non-rRNA Kraken2 clade counts for four obligately anaerobic human gut bacterial species: *Faecalibacterium prausnitzii, Roseburia intestinalis, Phocaeicola dorei*, and *Phocaeicola vulgatus*. These species are abundant core members of the healthy human gut microbiome that cannot proliferate in aerobic sewer infrastructure, making them a specific indicator of recent human fecal input decoupled from resident sewer biofilms (Newton et al., 2015; Roguet et al., 2022).

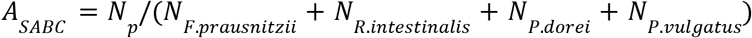

#### Time series alignment, pathogen matching, correlation, and significance testing

To compare WW-MGS, WW-PCR, and clinical time series, we first applied normalization at the sample level, and then aggregated data to MMWR epidemiological weeks. When multiple WW-MGS or WW-PCR samples were collected in a single MMWR week, we averaged values within that week using the geometric mean; clinical data were provided in weekly format and required no within-week aggregation. For this retrospective evaluation, we then applied a centered 5-week moving average to each time series independently using all available weeks for that data source. For WW-MGS and WW-PCR series, we smoothed values using the geometric mean over the 5-week window, computed as expm1(mean(log1p(x))) to remain defined when zero-valued observations enter the window. For Cosmos and NHSN clinical encounter counts, we used an arithmetic mean over the 5-week window, consistent with the conventional treatment of Poisson-distributed count data. We smoothed each series independently before restricting to the overlap window, ensuring that weeks near the edges of the overlap period benefited from a full smoothing window when one data source extended further in time than the other. We plotted smoothed values at the MMWR week midpoint, Wednesday.

For each pathogen, we matched the WW-MGS NCBI taxonomic identifiers (taxids) to the specificity of the corresponding PCR assay or clinical data. For WW-PCR comparisons, we matched WW-MGS taxids to the target specificity of each assay. For example, the WastewaterSCAN norovirus assay targets the ORF1-ORF2 junction of genogroup II specifically, so WW-MGS comparisons against WW-PCR used norovirus GII reads only. For the clinical norovirus comparison, we used the broader Norwalk virus taxid, capturing genogroups GI, GII, and GIV. For influenza, we distinguished between influenza A and B where possible, using type-specific WW-PCR data and clinical laboratory-confirmed tests from Cosmos. We also compared general influenza clinical encounters from NHSN HRD and Cosmos’ ICD-10 J09–J11 encounters with a combined influenza A and B WW-MGS signal. Supplementary Table S2 provides full taxid mappings for all pathogens and comparison sources.

We quantified concordance between WW-MGS, WW-PCR, and clinical trends using Spearman rank correlation coefficients computed on the smoothed values for MMWR weeks with observations from both sources. We chose Spearman correlation for its robustness to non-linear monotonic relationships and because it does not assume normality. We quantified pairwise relationships between normalization markers using Pearson correlation on log_10_-transformed marker fractions. We required at least ten overlapping MMWR weeks between the WW-MGS signal and the comparison signal for a correlation to be reported; within those overlapping weeks, we additionally required at least two MMWR weeks with non-zero observations on each side. We excluded site × pathogen × source combinations failing either criterion from analysis. Supplementary Figure S6 reports the sensitivity of the headline ΔR to the choice of smoothing window, including raw values without smoothing, the minimum overlap-weeks threshold, and the minimum non-zero comparison-weeks threshold.

To test whether normalization changed concordance with comparison signals, we computed ΔR as the change in Spearman R from the baseline total read relative abundance (A_total_) to each normalized abundance, paired within each site. We matched statistical inference on ΔR to each source’s comparison-side data structure: WW-PCR cells used per-site analysis because every site has its own sewershed PCR comparison series, while Cosmos and NHSN HRD cells used state-level analysis because all sites within a state share a single state-aggregated comparison series. For each normalization × pathogen × source combination, we applied a two-sided Wilcoxon signed-rank test to per-site ΔR values for WW-PCR or per-state mean ΔR values for Cosmos and NHSN, against the null hypothesis that the median was zero. Tests required at least six effective units, defined as sites for WW-PCR, states for Cosmos and NHSN, to be reported. This n ≥ 6 floor ensures that p < 0.05 is achievable; at n = 5, the smallest possible two-sided Wilcoxon p-value is 0.0625, regardless of effect size or directional consistency. This limit applies in particular to WW-PCR normalization analyses available only at WastewaterSCAN sites, including influenza B, HMPV, rotavirus, norovirus, EV-D68; n = 5 sites, which are reported descriptively without a significance test.

Within each pathogen × source combination, we adjusted p-values for multiple comparisons across the seven normalization methods using the Benjamini–Hochberg procedure with a false discovery rate of α = 0.05. Significance levels (*p < 0.05, **p < 0.01, ***p < 0.001) and significance counts reported throughout refer to Benjamini–Hochberg-adjusted p-values; the direction of effect is given by the sign of the median ΔR. We summarized uncertainty in the median ΔR using percentile 95% confidence intervals from 10,000 bootstrap resamples: per-site resampling for WW-PCR cells and a cluster bootstrap that resampled states with replacement, preserving within-state clustering, for Cosmos and NHSN cells. Supplementary Table S4 reports tabular results.

## Results

### Compositional distortion motivates wastewater metagenomic sequencing (WW-MGS) data normalization

Sequencing returns a fixed number of reads per sample, so changes in any one taxon’s abundance can distort the apparent abundance of every other taxon, even when their underlying absolute abundances are unchanged (Figure 1a–b). In the WW-MGS data, rRNA constituted a median of 61.0% of reads across all samples (interquartile range (IQR) 47.1–77.8%, n = 1,425) and varied substantially over time within sites, making it a dominant source of compositional variation in the sequencing libraries (Supplementary Figure S7). Variation in rRNA fraction propagated to all non-rRNA taxa, including normalization markers; at Boston Deer Island Treatment Plant North (DITPN), PMMoV fraction was strongly negatively correlated with rRNA fraction (Spearman R = −0.79, p = 4.3 × 10^−17^, n = 76; Figure 1c–d). SARS-CoV-2 total read relative abundance showed no correlation with Massachusetts Cosmos clinical encounters (R = 0.10, p = 0.37; Figure 1e), but PMMoV normalization recovered concordance (R = 0.74, p = 1.6 × 10^−14^; Figure 1f), consistent with the interpretation that compositional variation in the sequencing library was obscuring the underlying pathogen trend.

**Figure 1.**
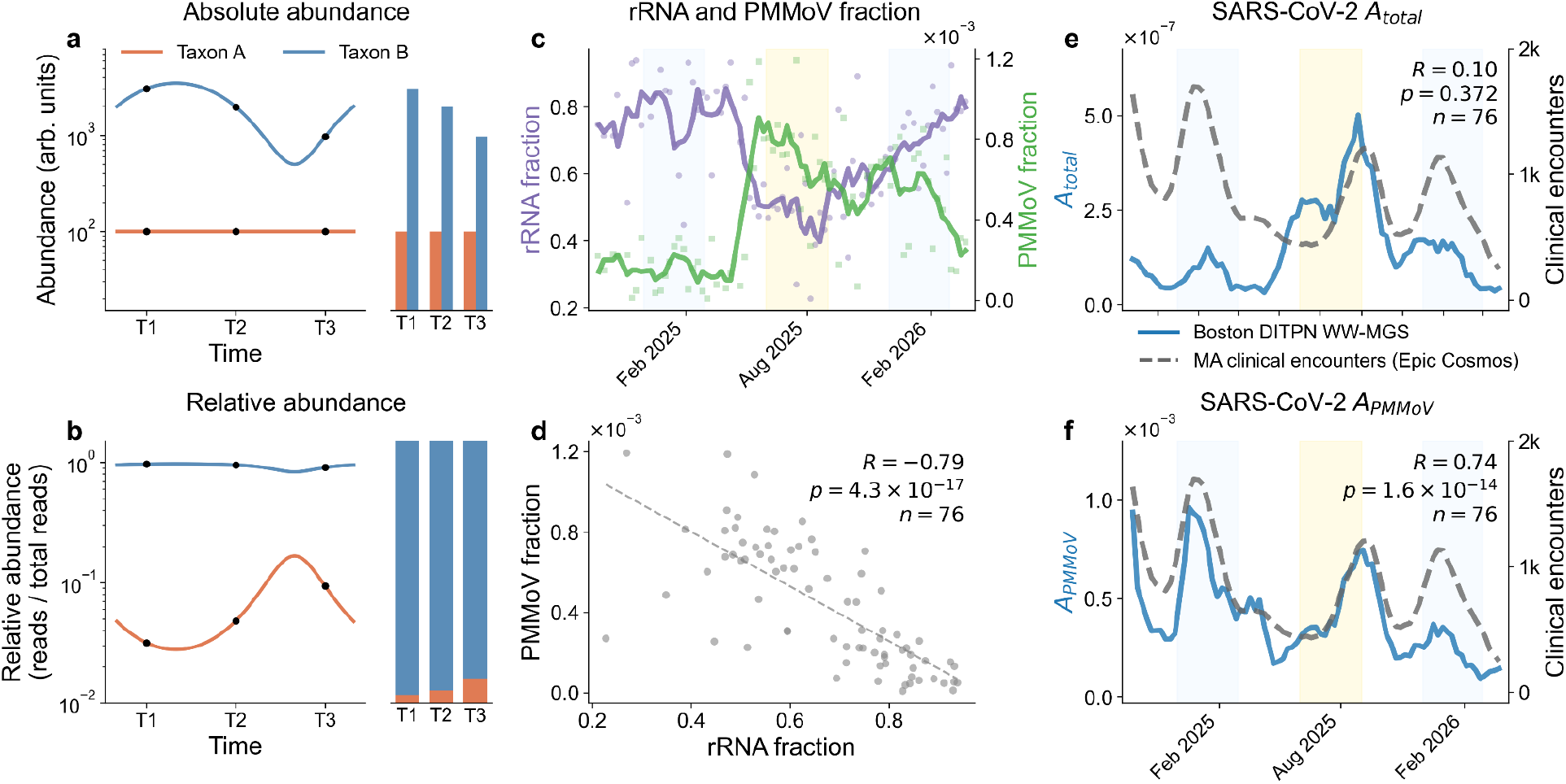
Compositional distortion of apparent pathogen relative abundance in wastewater metagenomic sequencing (WW-MGS). (a) Schematic example showing the absolute abundance of two taxa over time, as would be measured by quantitative PCR (qPCR). (b) The same two taxa in a compositional sequencing framework, where each taxon is expressed as a fraction of total reads; fluctuations in Taxon B distort the apparent abundance of the otherwise-constant Taxon A. (c) Fraction of ribosomal RNA (rRNA) and pepper mild mottle virus (PMMoV) reads over time at the Boston Deer Island Treatment Plant North (DITPN) site. (d) PMMoV fraction versus rRNA fraction at Boston DITPN. (e) SARS-CoV-2 total read relative abundance (A_total_) at Boston DITPN compared to Epic Cosmos (Cosmos) state-level clinical encounters. (f) PMMoV-normalized SARS-CoV-2 abundance (A_PMMoV_) at the same site compared to Cosmos.

### Candidate normalization markers differ in abundance and stability

The utility of a normalization marker depends on its abundance, temporal stability, and biological specificity for the compositional variation it is intended to control. Low-abundance markers can add sampling noise to the denominator, while markers with strong seasonal trends can introduce new compositional artifacts. Candidate markers also track biologically distinct signals, ranging from diet-derived fecal indicators to endogenous gut microbes to overall library composition, and it is uncertain *a priori* which signal best reflects human fecal strength and pathogen shedding. We therefore characterized all seven normalization candidates before evaluating their effects on concordance (Figure 2).

**Figure 2.**
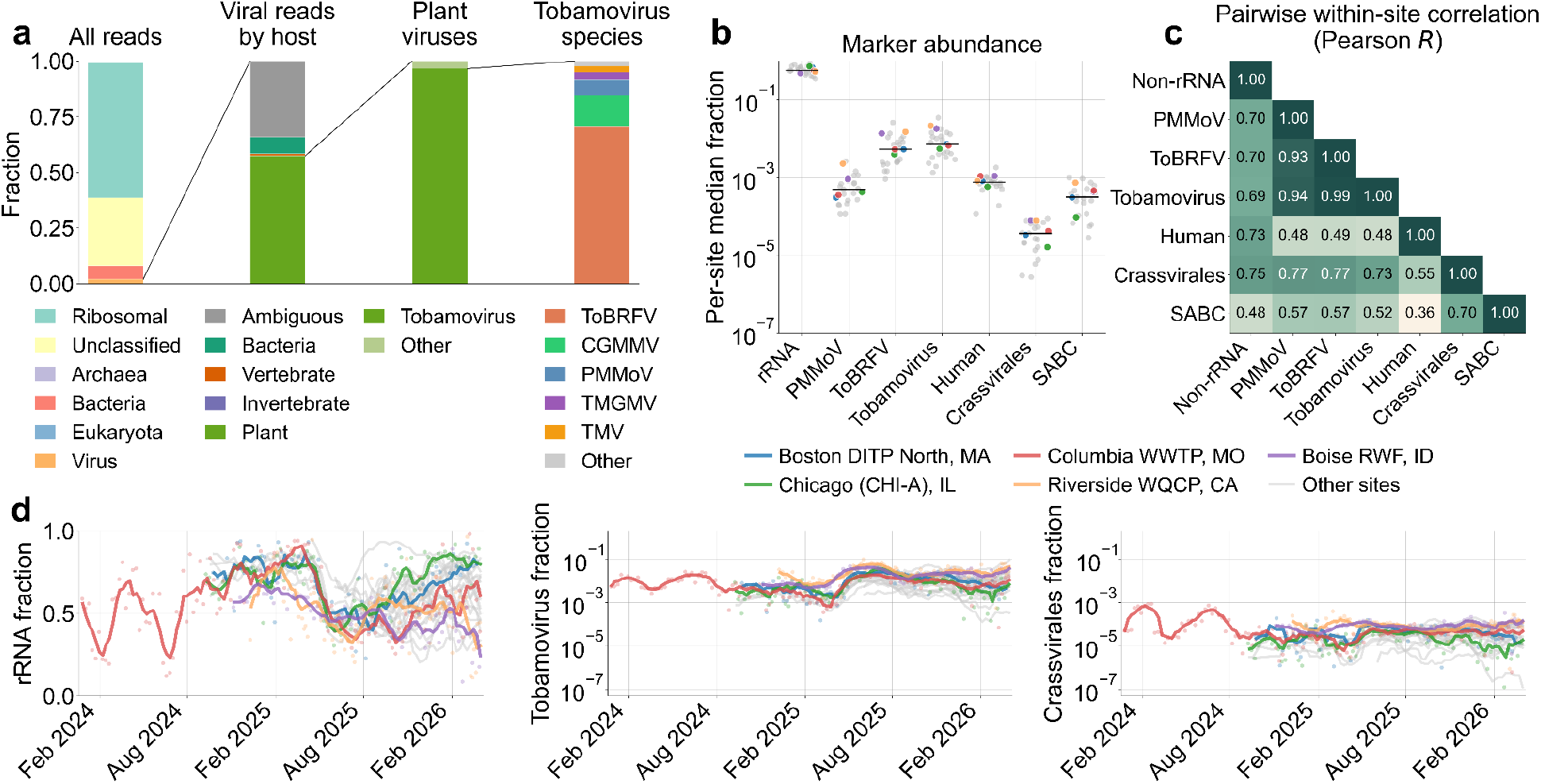
Wastewater metagenomic sequencing (WW-MGS) normalization marker abundance and dynamics. (a) Aggregate taxonomic composition showing all reads split by domain, viral reads by host, plant viruses by *Tobamovirus* versus other genera, and *Tobamovirus* reads by species. (b) Per-site median marker fractions; each point represents one site (n = 25), the horizontal black bar marks the across-site median, and the five highlighted sites are colored as in (d). (c) Pairwise within-site Pearson R on log_10_-transformed marker fractions. Each site’s mean log_10_(marker) is subtracted before pooling, so correlations refl ect sample-to-sample co-variation within sites. (d) Morbidity and Mortality Weekly Report (MMWR)-smoothed temporal trends of ribosomal RNA (rRNA), tomato brown rugose fruit virus (ToBRFV), and *Crassvirales* fractions. ToBRFV and Crassvirales are shown with log-scaled y-axes. Five highlighted sites are shown in color, and other sites are shown in gray. CGMMV, cucumber green mottle mosaic virus; PMMoV, pepper mild mottle virus; TMGMV, tobacco mild green mosaic virus; TMV, tomato mosaic virus.

Tobamoviruses dominated the Kraken2-classified viral community, making up a median of 53% of viral reads (IQR 39.0–65.1%) across samples (Supplementary Figure S8). Among tobamoviruses, ToBRFV was the most abundant species, accounting for a median of 72% of genus-level reads, and was roughly 10-fold more abundant than PMMoV (median ratio 10.1×, IQR 6.8–14.9×). Per-site relative abundances of these markers were generally consistent over multi-month scales (Figure 2d, Supplementary Figure S9), although some tobamovirus species, such as cucumber green mottle mosaic virus, tomato mosaic virus, and ToBRFV showed signs of seasonal abundance trends (Supplementary Figure S10-11). Endogenous gut markers were 10-to 250-fold less abundant than tobamovirus markers, suggesting that they may introduce noisier denominators when used for normalization. Pairwise log-Pearson correlations clustered the diet-derived tobamoviruses tightly (PMMoV–ToBRFV R = 0.93, ToBRFV–tobamovirus R = 0.99; p < 10^−300^; Figure 2c, Supplementary Figure S12), consistent with similar normalization performance across these markers. Endogenous gut markers correlated more strongly with one another than with tobamoviruses (Figure 2c).

### Normalization improves concordance with established epidemiological signals

We evaluated seven normalization approaches against the total read relative abundance baseline (A_total_) for eight pathogens across 25 sites, comparing against WW-PCR from WastewaterSCAN and NWSS, Cosmos clinical encounters, and NHSN HRD admissions (Figure 3).

**Figure 3.**
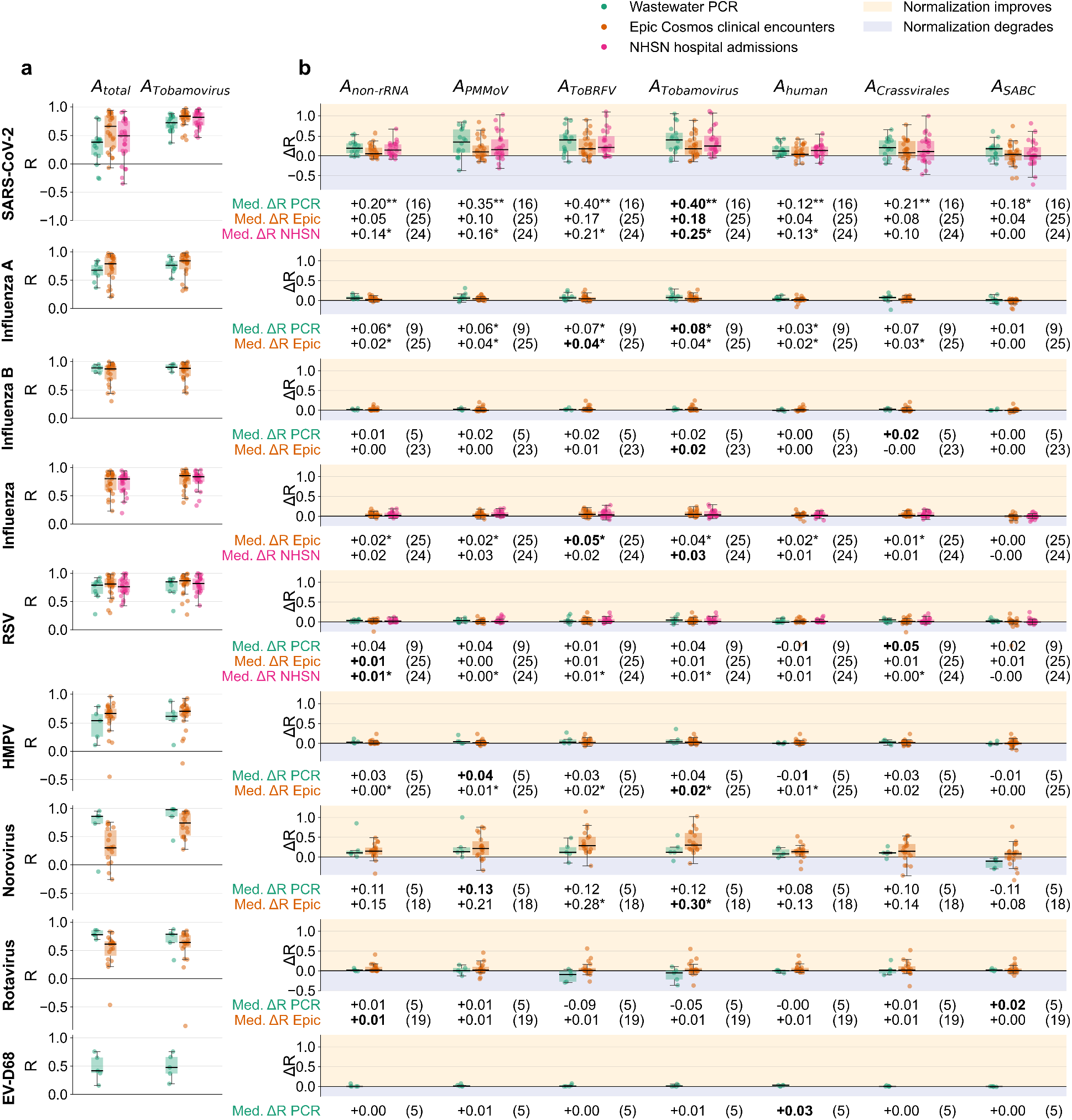
Concordance and per-normalization method ΔR for wastewater metagenomic sequencing (WW-MGS) against wastewater PCR (WW-PCR) and clinical comparison sources. (a) Per-site Spearman R between WW-MGS total read relative abundance (A_total_, left) and tobamovirus genus-normalized abundance (A_Tobamovirus_, right) against each comparison source. Colored boxplots show WW-PCR, Epic Cosmos clinical encounters, National Healthcare Safety Network (NHSN) hospital admissions; each dot represents one site. (b) Paired ΔR (R_normalized_ − R_A_total_) across seven normalization methods for the same pathogen × source combinations. Numbers below each boxplot indicate the median ΔR with significance stars from Benjamini–Hochberg-adjusted two-sided Wilcoxon signed-rank test against the null hypothesis of median ΔR = 0 (*, p < 0.05; **, p < 0.01; ***, p < 0.001). Parentheses indicate the number of sites contributing to each pathogen × source pairing. The “Infl uenza (combined)” row represents combined WW-MGS infl uenza A and B relative abundance compared with NHSN’s combined influenza admissions and Cosmos’ International Classification of Diseases, 10th Revision (ICD-10) J09–J11 general-infl uenza encounters.

Baseline A_total_ concordance varied widely across pathogens (Figure 3a). Median Spearman R against Cosmos clinical encounters ranged from 0.30 for norovirus to 0.87 for influenza B, and median Spearman R against WW-PCR ranged from 0.38 for SARS-CoV-2 to 0.89 for influenza B. Influenza and RSV, which show distinct winter peaks separated by periods of absence, reached higher baseline correlations than the year-round-circulating pathogens such as SARS-CoV-2 and norovirus. Norovirus had a strong baseline concordance with WW-PCR (R=0.86) but the lowest concordance with Cosmos (R=0.30) of any pathogen, reflecting sparser clinical signal (~16,000 Cosmos encounters across included states during the sampling window, versus >100,000 for SARS-CoV-2, influenza, and RSV; Supplementary Figure S4). HMPV and rotavirus had intermediate baseline A_total_ concordance with Cosmos (R = 0.67 and 0.61) on similarly limited clinical data (~17,000 and ~1,800 encounters), and together with EV-D68 were matched to only five WW-PCR sites, making their concordance estimates less reliable.

Normalization improved concordance for most pathogen × comparison source combinations (Figure 3b), with benefit varying strongly by pathogen. SARS-CoV-2 and norovirus, which started from low baselines, showed the largest gains. Tobamovirus-genus normalization increased median R against WW-PCR for SARS-CoV-2 from 0.38 to 0.72 (median ΔR = +0.40, p = 0.0010, n = 16 sites) and increased median R against Cosmos for norovirus from 0.30 to 0.73 (median ΔR = +0.30, p = 0.027, n = 18). Influenza and RSV showed small but generally significant gains, consistent with ceiling effects limiting gains at higher baselines. HMPV showed only a marginal but significant benefit with Cosmos (ΔR = +0.021, p = 0.027) and was underpowered against WW-PCR (n = 5 sites); EV-D68 had no comparator with sufficient power for significance testing. Rotavirus was the only pathogen for which tobamovirus normalization reduced median concordance against WW-PCR (median ΔR = −0.054, n = 5 sites; underpowered for significance testing).

The three tobamovirus-based markers (PMMoV, ToBRFV, and tobamovirus genus) generally performed best among normalization approaches and similarly to one another, consistent with their tight correlation (Figure 2c). PMMoV-normalized abundance underperformed ToBRFV and tobamovirus-genus normalization for SARS-CoV-2 across all comparison sources, potentially reflecting PMMoV’s ~10-fold lower abundance. Non-rRNA normalization improved over baseline but trailed the fecal-indicator viruses. Endogenous gut markers performed less consistently than tobamovirus markers, with SABC-normalized abundance producing no significant gain for any tested combination. In the next section, we focus on tobamovirus genus normalization.

### Tobamovirus genus normalization demonstrates strong performance across pathogens and comparators

Tobamovirus genus normalization improved concordance against clinical and WW-PCR signals across nearly all pathogens and rarely degraded performance. Median ΔR was positive for 18 of 19 pathogen × comparison-source combinations, with bootstrapped 95% confidence intervals excluding zero in 10 of the 14 combinations meeting the inference floor (Figure 4a). SARS-CoV-2 and norovirus showed the largest gains. RSV, influenza, and HMPV, which are defined by sharp winter peaks, also showed small but consistent benefits, consistent with ceiling effects from higher A_total_ baselines (Figure 4a). For rotavirus, tobamovirus genus normalization was inconsistent against Cosmos and slightly reduced median concordance with WW-PCR, where the analysis was limited to five sites.

**Figure 4.**
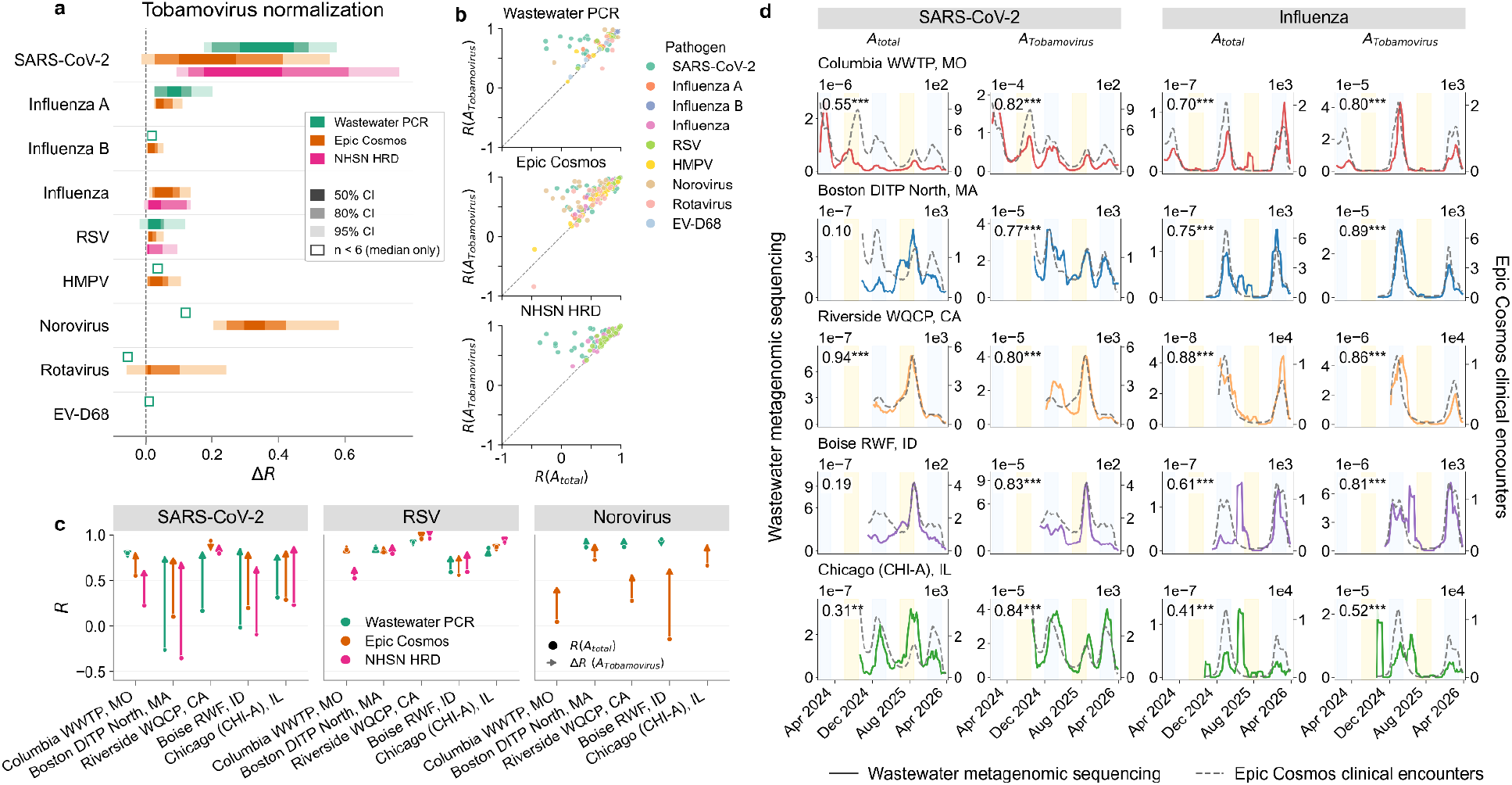
Impact of tobamovirus genus normalization on concordance with epidemiological signals across pathogens, sites, and comparison sources. (a) Median ΔR (tobamovirus-normalized A_Tobamovirus_ - A_total_) by pathogen, stratified by comparison source: wastewater PCR (WW-PCR), Epic Cosmos clinical encounters, and National Healthcare Safety Network (NHSN) hospital admissions. Bars show bootstrap quantile intervals from 10,000 resamples on the median ΔR across sites. (b) Spearman R under tobamovirus genus normalization versus baseline A_total_ for every site × pathogen combination, shown in separate panels by comparison source. Points above the y = x diagonal indicate combinations where tobamovirus genus normalization improved concordance; points below the diagonal indicate combinations where it reduced concordance. (c) Per-site change in Spearman R from baseline A_total_ to tobamovirus-normalized A_Tobamovirus_ for SARS-CoV-2, RSV, and norovirus at five representative sites. (d) Five-week smoothed time series of SARS-CoV-2 and influenza at the same five sites, comparing baseline A_total_ and tobamovirus genus-normalized abundance against Epic Cosmos clinical encounters. Spearman R is annotated in each panel; significance is indicated as *p < 0.05, **p < 0.01, and ***p < 0.001. Background shading indicates winter (blue) and summer (yellow) seasons.

The magnitude of ΔR was asymmetric in both direction and magnitude (Figure 4b). Across 316 site × pathogen × comparator combinations, 246 (78%) had positive ΔR and 62 (20%) had negative ΔR. When normalization improved concordance, the medial gain was +0.09, approximately 5× larger than the median loss when normalization reduced concordance (−0.02).

The impact of normalization was largely consistent across comparison sources (Figure 4c). Among 98 site × pathogen combinations with paired data from two or more comparators, 81 (83%) had all non-zero ΔR values in the same direction (Supplementary Figure S13).

In contrast to WW-MGS, where tobamovirus genus normalization consistently improved concordance across pathogens and comparison sources, PMMoV normalization of WW-PCR had mixed and source-dependent effects (Supplementary Figure S2, Supplementary Table S5). At WastewaterSCAN sites, the median per-site × pathogen ΔR from PMMoV normalization was near zero against both Epic Cosmos encounters (median ΔR = −0.007, n = 31) and NHSN admissions (median ΔR = −0.010, n = 20). At NWSS sites with PMMoV normalization available, limited here to the four Chicago sites, PMMoV normalization shifted concordance by median ΔR = −0.13 against Epic Cosmos (n = 12) and +0.03 against NHSN (n = 12) for the same SARS-CoV-2, influenza A, and RSV measurements. This inconsistent normalization impact is consistent with mixed reports in the WW-PCR literature on the value of fecal-indicator normalization for WW-PCR data (Dhiyebi et al., 2023; Duvallet et al., 2022; Hsu et al., 2022; Maal-Bared et al., 2023).

Tobamovirus genus-normalized WW-MGS reached concordance with clinical comparators on par with WW-PCR (Supplementary Figure S14). For SARS-CoV-2, median Spearman R between A_Tobamovirus_ and Epic Cosmos data was 0.83 (n = 25 WW-MGS sites), slightly higher than raw WW-PCR concordance with Cosmos (0.72, n = 16 WW-PCR sites) and PMMoV-normalized WW-PCR (0.75, n = 9). WW-MGS and raw WW-PCR showed similar concordance with Cosmos for Influenza A and RSV (Influenza A: 0.84 vs 0.86; RSV: 0.87 vs 0.86) and with NHSN admissions for SARS-CoV-2 (0.81 vs 0.70) and RSV (0.82 vs 0.82).

## Discussion

WW-MGS is a powerful emerging tool for quantitative monitoring of hundreds of known and potentially novel pathogens across human, animal, and plant populations. However, the naive approach to tracking pathogen abundance in WW-MGS data, total read relative abundance, is prone to biases that can distort the measured signal away from the underlying epidemiological dynamics it is meant to track. These biases arise from variation in sample processing and recovery, which WW-MGS shares with WW-PCR, and from the compositional structure of sequencing data itself, where a taxon’s apparent abundance depends on the abundance of every other taxon in the library. In this retrospective analysis, we show that normalizing WW-MGS abundance reliably improves concordance with established epidemiological signals, including clinical encounters and WW-PCR. Among the seven normalization approaches we evaluated, tobamovirus-based markers, which draw on a genus of plant viruses abundant in human stool, performed best.

The benefits of normalization varied by pathogen, were directionally consistent across comparison sources, and generally outweighed the cases where normalization reduced concordance. SARS-CoV-2 and norovirus, pathogens with substantial year-round circulation, showed the largest gains, consistent with year-round exposure to compositional distortion as the abundance of other taxa in the background fluctuates. Sharply seasonal pathogens such as influenza and RSV had higher baseline correlations, with concordance dominated by periods of presence and absence, leaving less room for normalization to improve correlation. The direction of normalization effects was largely consistent across WW-PCR, clinical, and hospitalization comparators, supporting the interpretation that gains reflect recovery of true epidemiological signal rather than artifacts of any single comparator. When normalization degrades concordance, the losses were generally small. Tobamovirus genus-normalized abundance produced median gains roughly fivefold larger than median losses across all pathogens and sources, an asymmetry that makes tobamovirus genus a low-risk default for operational WW-MGS surveillance when pathogen-specific normalization data are not available. The magnitude of normalization benefit also varied across sites (Figure 4), perhaps because the compositional and processing variation that normalization corrects is itself site-specific. Monitoring networks may therefore benefit from verifying normalization performance per site as deployments expand.

The strong performance of normalization in WW-MGS contrasts with mixed results in the WW-PCR literature, where PMMoV and flow-based normalization improve concordance with clinical data at some sites but not others (Dhiyebi et al., 2023; Duvallet et al., 2022; Hsu et al., 2022; Maal-Bared et al., 2023). Part of this inconsistency likely reflects heterogeneity in upstream sample processing across WW-PCR studies, with concentration and extraction methods varying widely between WW-PCR programs and serving as major drivers of measured marker variability (Pappu et al., 2026). Nevertheless, the contrast between WW-PCR and WW-MGS normalization helps localize where gains in WW-MGS come from. WW-PCR and WW-MGS share biases related to sample collection and processing, but WW-MGS is also affected by the compositional structure of sequencing, where any taxon’s apparent abundance depends on the abundance of every other taxon sequenced alongside it. In our analysis, comparing raw WW-PCR concentrations with PMMoV- and flow-normalized WW-PCR produced little change in concordance with clinical comparators, whereas applying the same fecal-indicator markers to WW-MGS produced large and consistent gains. This pattern suggests that fecal-indicator normalization in WW-MGS primarily corrects sequencing-specific compositional distortion rather than biases shared with WW-PCR.

A broader implication of these results is that normalized WW-MGS achieves concordance with clinical epidemiological signals similar to that of targeted WW-PCR for the same pathogens. WW-MGS has been positioned primarily as a discovery tool, well suited to pathogen-agnostic detection of both known and novel agents. With appropriate normalization, WW-MGS also functions as a quantitative trend-tracking tool, with the additional advantage that a single sequencing assay yields data for every pathogen detectable in the library without requiring separate validated PCR assays for each target. As WW-MGS becomes a more standard component of pathogen surveillance infrastructure, treating it as a quantitative monitoring platform substantially expands its value beyond detection alone.

One limitation of this analysis is that tobamovirus genus normalization may not generalize uniformly across geography, time, or wastewater systems. All included CASPER sites are in the continental U.S., and while tobamoviruses are abundant in the global wastewater virome (Symonds et al., 2018; Worp et al., 2025), regional variation in diet, agriculture, and food supply chains could shift the relative abundance and temporal dynamics of individual species (Pappu et al., 2026). Consistent with this, several tobamovirus species, including ToBRFV, showed signs of seasonal abundance trends in our data that may introduce bias into the denominator independent of human fecal input strength (Supplementary Figure S10). The recent global spread of ToBRFV through commercial tomato production may also make its wastewater abundance especially sensitive to future changes in agricultural prevalence or control. The compositional structure of WW-MGS makes it difficult to attribute such trends to changes in a marker’s absolute shedding rather than to shifts in the surrounding library, but seasonal variation in diet and agricultural inputs is a plausible contributor and warrants further investigation. Still, tobamovirus normalization empirically improved concordance across pathogens despite these trends, and normalizing by the tobamovirus genus rather than relying on any single species may reduce dependence on the seasonal, regional, or agricultural dynamics of any one marker.

Other limitations arise from the structure of the evaluation itself. The per-site ΔR framework, which measures the change in Spearman correlation between total read relative abundance and the normalized WW-MGS signal relative to a given comparator, collapses each site’s time series into a single number and weights all sites equally regardless of time series length. The metric also ignores features such as peak timing or growth-rate estimation, which are epidemiologically relevant parameters. The retrospective smoothing strategy also limits direct operational interpretation. We used centered 5-week moving averages to compare noisy time series, but a real-time system cannot use future observations to smooth the current week. Operational deployment would require causal approaches, such as trailing-window smoothing, state-space or nowcasting models, or anomaly-detection methods that update estimates as new WW-MGS data arrive. Finally, the framework implicitly treats WW-PCR, clinical encounters, and hospital admissions as closer approximations of pathogen burden than WW-MGS, which is not necessarily the case (Ahmed et al., 2026). Clinical encounter and hospitalization signals are themselves biased toward symptomatic individuals seeking care and miss the asymptomatic or subclinical infections that wastewater is well-positioned to capture. As a result, perfect correlation with these comparators is neither realistic nor desirable.

As WW-MGS deployments expand, quantitative interpretation of pathogen trends will require standard practices for handling compositional distortion. Tobamovirus genus normalization in U.S. settings provides a strong empirical foundation for those practices. However, we evaluated only a fixed set of candidate markers and did not systematically scan all possible markers or combine markers into composite denominators that might further reduce noise, so stronger approaches may exist. Future work should explore this larger space of markers and denominators, evaluate additional comparison data sources and concordance metrics, extend normalization to hybrid-capture WW-MGS (Tisza et al., 2023; Wolfe et al., 2026), where the compositional structure differs from untargeted sequencing, pursue absolute quantification with spike-in controls (Hardwick et al., 2018; Langenfeld et al., 2025), and test the global representativeness of these findings. Together, these next steps can help turn WW-MGS from a pathogen-agnostic detection method into a standardized quantitative surveillance platform

## Data availability

WW-MGS abundance data and the comparison-source time series used in this analysis, together with code to reproduce all results, are available at https://github.com/lennijusten/wastewater-sequencing-normalization. Raw CASPER sequencing data are available under NCBI BioProject accession PRJNA1247874, and the CASPER WW-MGS dataset is described in Justen et al. (2026). Prior to public release on SRA, human-derived reads were masked using NCBI’s Human Read Removal Tool. WW-PCR data are from WastewaterSCAN (Boehm et al., 2026) and the CDC NWSS (CDC, 2025a, 2025b, 2025c). WastewaterSCAN data are shared under a CC BY-NC 4.0 license; see https://data.wastewaterscan.org/about for terms of use. WastewaterSCAN data were collected as part of the WastewaterSCAN / SCAN project, a partnership between Stanford University, Emory University, and Verily funded philanthropically through a gift to Stanford University. Clinical encounter data are from the Epic Cosmos dataset (Epic Systems Corporation, Verona, WI) and were obtained through an institutional agreement. Data are available at https://cosmos.epic.com/. Hospital admission data are from the CDC National Healthcare Safety Network Hospital Respiratory Data (CDC, 2024).

## Funding

L.J.J. was supported by the Draper Scholar program at The Charles Stark Draper Laboratory.

## Acknowledgements

We thank Elana Chan and Daniel Rice for helpful feedback on the manuscript, and Mike Tisza for useful input in various communications. We are grateful to the members of the CASPER network for wastewater sample collection and processing, and to the SecureBio Detection team for development of the bioinformatic infrastructure underlying metagenomic sequencing taxonomic classification. Work by RSK was performed under the auspices of the U.S. Department of Energy by Lawrence Livermore National Laboratory under Contract DE-AC52-07NA27344.

## Author contributions

**Conceptualization:** L.J., M.R.M. **Methodology:** L.J., R.Y.L., M.R.M. **Software:** L.J. **Validation:** L.J. **Formal analysis:** L.J. **Investigation:** L.J. **Resources:** L.J., R.Y.L., L.S.M., J.K., M.C.J., P.C.S. **Data curation:** L.J. **Writing – original draft:** L.J. **Writing – review & editing:** L.J., A.Z., R.S.K., R.Y.L., D.C.-B., J.K., M.C.J., M.R.M., P.C.S. **Visualization:** L.J. **Supervision:** D.C.-B., J.K., M.C.J., M.R.M., P.C.S. **Project administration:** L.J. **Funding acquisition:** J.K., M.C.J., P.C.S.

## Competing interests

P.C.S. holds several patents related to diagnostic and surveillance technologies and is a co-founder and equity holder in Delve Biosciences and Lyra Labs, a board member and equity holder in Polaris Genomics, and an equity holder of NextGenJane. P.C.S. was formerly a co-founder of Sherlock Biosciences and board member of Danaher Corporation, until December 2024. All potential conflicts are managed in accordance with institutional policy.

## Notes

### Author Declarations

The Stanford University Institutional Review Board determined that this project does not meet the definition of human subject research (Protocol 87814); Epic Cosmos data are deidentified.

