## Supplementary material for "Establishing wastewater metagenomics as a quantitative pathogen monitoring tool with normalization": si

### Table of contents


**Figures**

| | | |
|---|---|---|
| Supplementary Figure S1. | Sequencing depth (total read pairs) over time by site. | 3 |
| Supplementary Figure S2. | Impact of wastewater PCR (WW-PCR) normalization on concordance with wastewater metagenomic sequencing (WW-MGS) and clinical data. | 4 |
| Supplementary Figure S3. | Sensitivity of Epic Cosmos per-site ΔR to suppressed-cell handling. | 5 |
| Supplementary Figure S4. | Time series abundance of wastewater PCR (WW-PCR) and clinical data. | 6 |
| Supplementary Figure S5. | Normalization marker abundance per sample and per site. | 7 |
| Supplementary Figure S6. | Sensitivity of $A_{Tobamovirus}$ median ΔR to smoothing-window width, non-zero-weeks filter, and minimum-overlap-weeks threshold. | 8 |
| Supplementary Figure S7. | Taxonomic composition over time by site. | 9 |
| Supplementary Figure S8. | Tobamovirus reads as a fraction of viral reads over time by site. | 10 |
| Supplementary Figure S9. | Time series of normalization marker fractions as a fraction of total reads and non-rRNA reads. | 11 |
| Supplementary Figure S10. | Within-site temporal stability of normalization-marker fractions. | 12 |
| Supplementary Figure S11. | Tobamovirus genus composition over time by site. | 13 |
| Supplementary Figure S12. | Pairwise correlations between normalization marker fractions across all samples. | 14 |
| Supplementary Figure S13. | Site-level impact of normalizing wastewater metagenomic sequencing data with tobamovirus genus counts against wastewater PCR and clinical trends across eight pathogens. | 15 |
| Supplementary Figure S14. | Cross-source agreement between wastewater metagenomic sequencing, wastewater PCR, and clinical comparators. | 16 |
| Supplementary Figure S15. | Per site pathogen abundance trends in wastewater metagenomic sequencing (WW-MGS) data. | 17 |

**Tables**

| | | |
|---|---|---|
| Supplementary Table S1. | Wastewater PCR (WW-PCR) data sources mapped to CASPER wastewater metagenomic sequencing (WW-MGS) sites. | 18 |
| Supplementary Table S2. | Taxonomic and case-definition matching between wastewater metagenomic sequencing (WW-MGS) and each comparison source. | 19 |
| Supplementary Table S3. | Zero denominator counts for each normalization approach. | 20 |
| Supplementary Table S4. | Normalization performance across comparison sources by pathogen. | 21 |
| Supplementary Table S5. | Per-pathogen wastewater PCR (WW-PCR) normalization summary. | 22 |

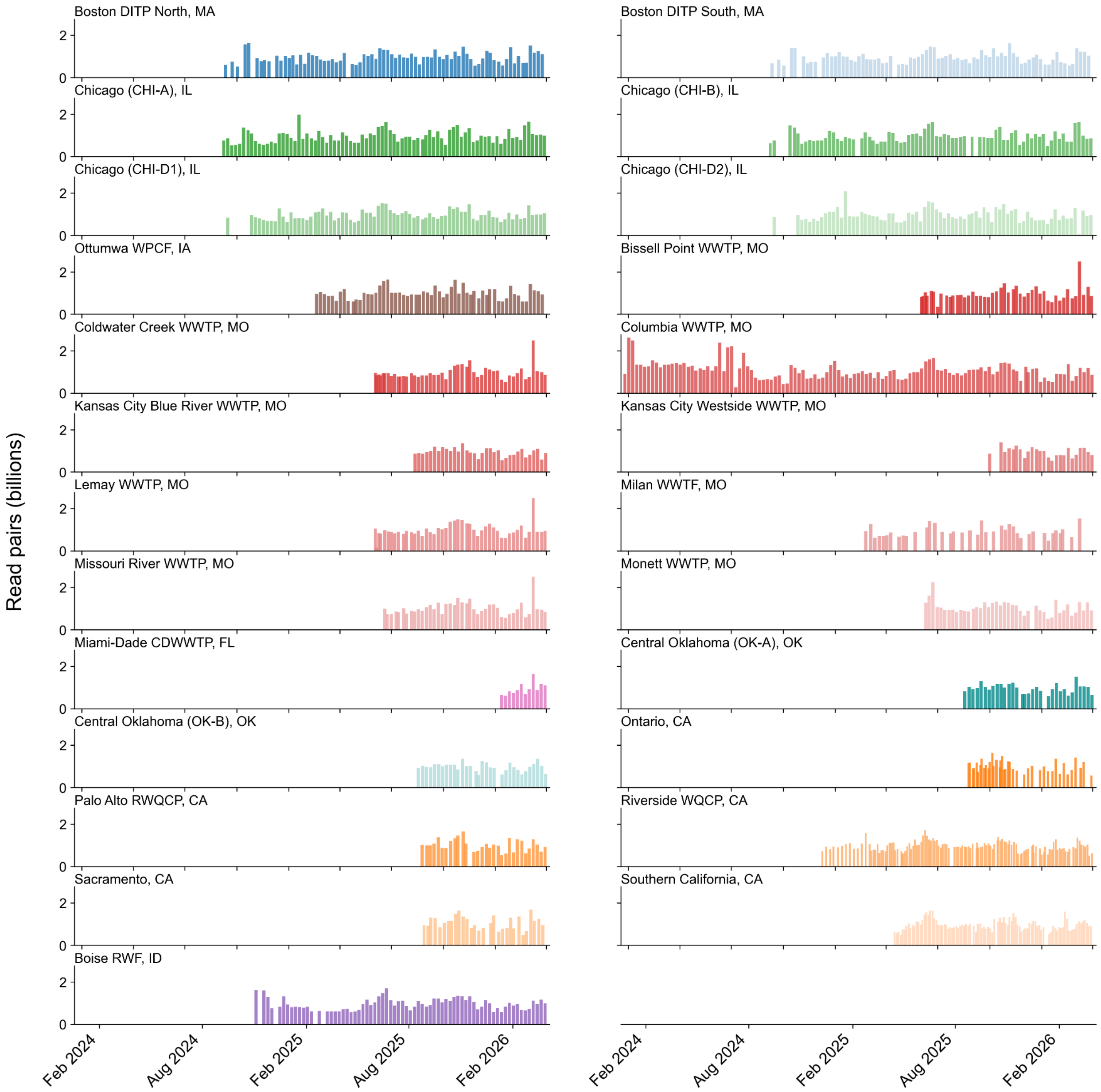


**Supplementary Figure S1. Sequencing depth (total read pairs) over time by site.**

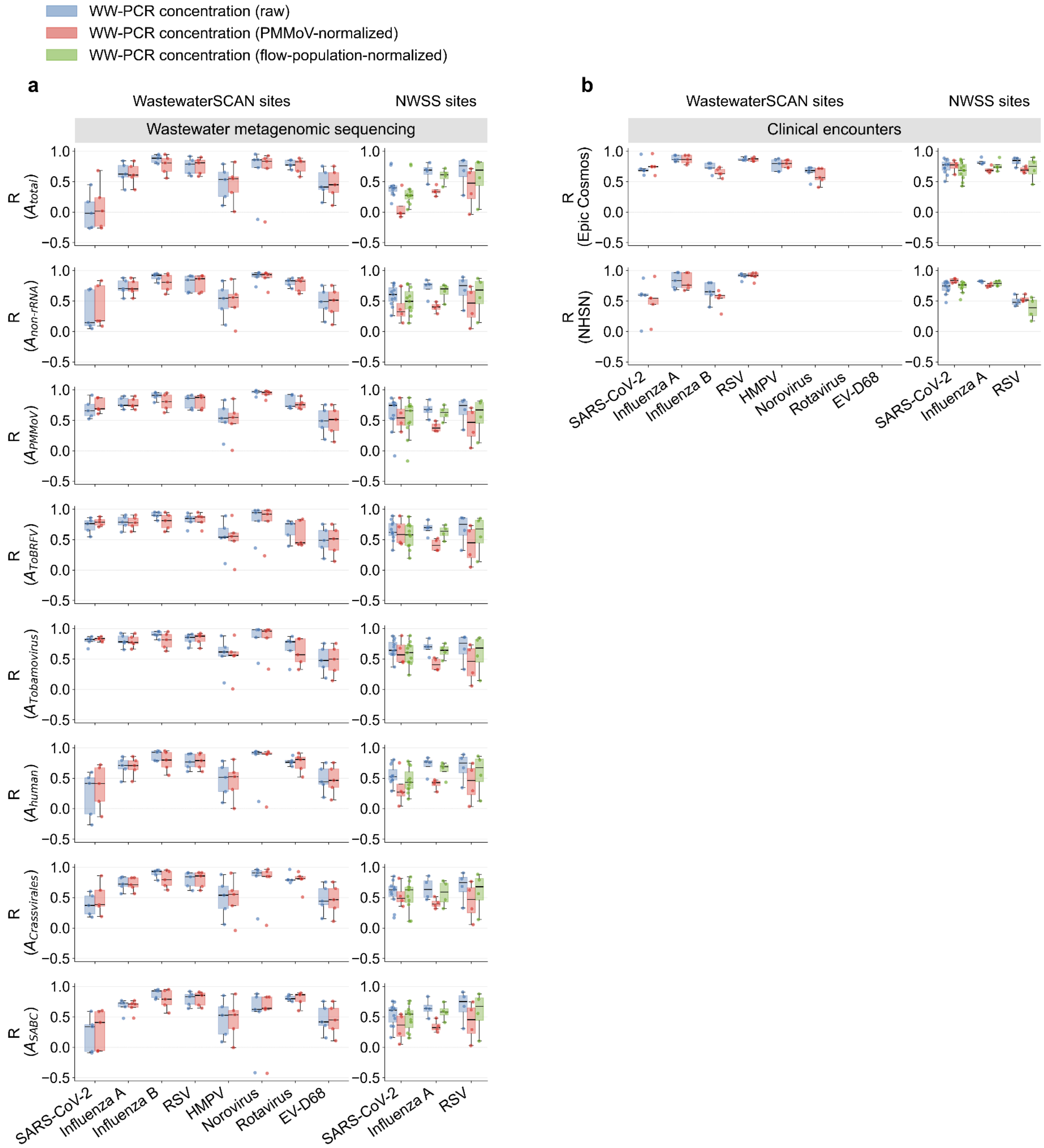


**Supplementary Figure S2. Impact of wastewater PCR (WW-PCR) normalization on concordance with wastewater metagenomic sequencing (WW-MGS) and clinical data.** Box plots show per-site Spearman R between WW-PCR concentration (raw or normalized) and the corresponding comparison signal; each dot is one site. (a) Comparisons against WW-MGS relative abundance under eight normalization methods, shown separately for WastewaterSCAN sites (left, ddRT-PCR on solids, gene copies (gc) per gram dry weight; (Boehm et al., 2026) and NWSS sites (right, qPCR or ddPCR on liquid, gc per liter; CDC NWSS data portal (Adams et al., 2024)). (b) Comparisons against state-level Epic Cosmos clinical encounters and National Healthcare Safety Network (NHSN) Hospital Respiratory Data (HRD) admissions, again split by PCR source. Wastewater PCR cells are restricted to the five WastewaterSCAN sites and eleven NWSS sites used in the main WW-MGS PCR analysis (Supplementary Table S1; WastewaterSCAN takes precedence at sites with both). PMMoV, pepper mild mottle virus; ToBRFV, tomato brown rugose fruit virus; SABC, strict-anaerobic-gut-bacteria composite.

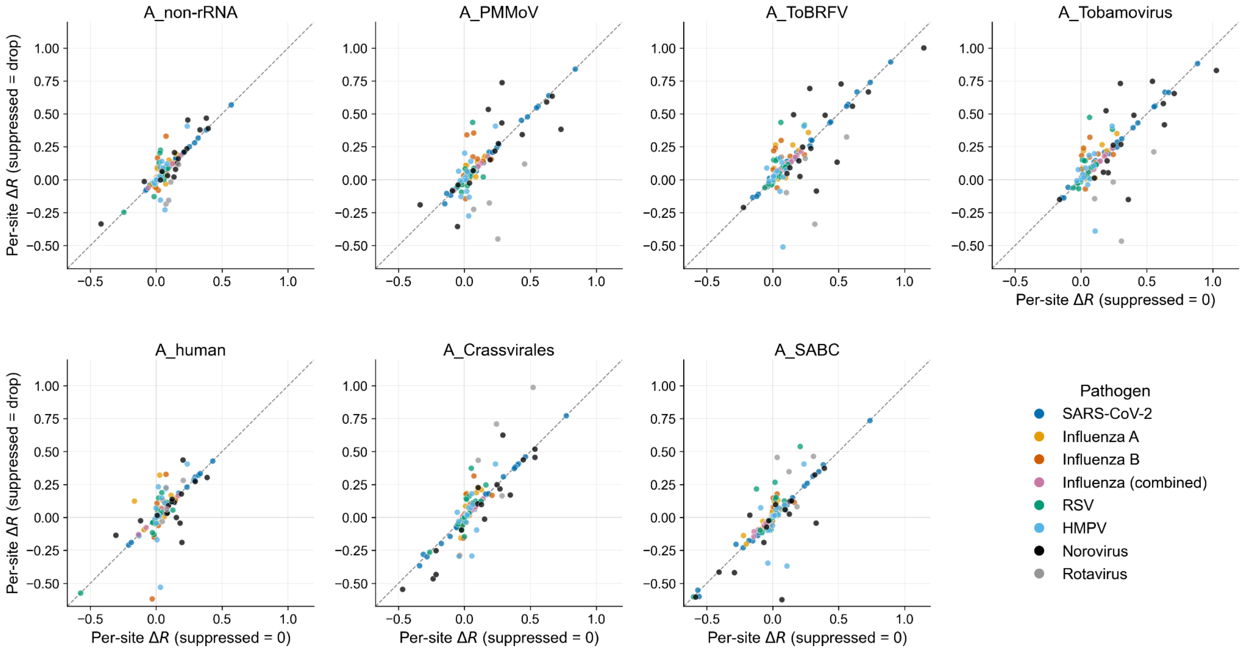


**Supplementary Figure S3. Sensitivity of Epic Cosmos per-site ΔR to suppressed-cell handling.** Epic Cosmos suppresses weekly encounter counts of ten or fewer to protect patient privacy. The main analysis treats suppressed cells as zero, preserving low-incidence weeks as informative observations; this figure compares that policy against dropping suppressed cells as missing values. Wastewater PCR (WW-PCR) and National Healthcare Safety Network (NHSN) Hospital Respiratory Data (HRD) comparisons are unaffected by this choice and not shown. Points indicate site-level ΔR values (the per-(site × pathogen) change in Spearman R relative to the total-read relative abundance baseline $A_{total}$), with the value under the suppressed = 0 policy and the value under the suppressed = drop policy. Sites are restricted to those passing the headline inclusion criterion (≥10 overlapping weeks and ≥2 non-zero weeks on each side) under both policies. PMMoV, pepper mild mottle virus; ToBRFV, tomato brown rugose fruit virus; SABC, strict-anaerobic-gut-bacteria composite.

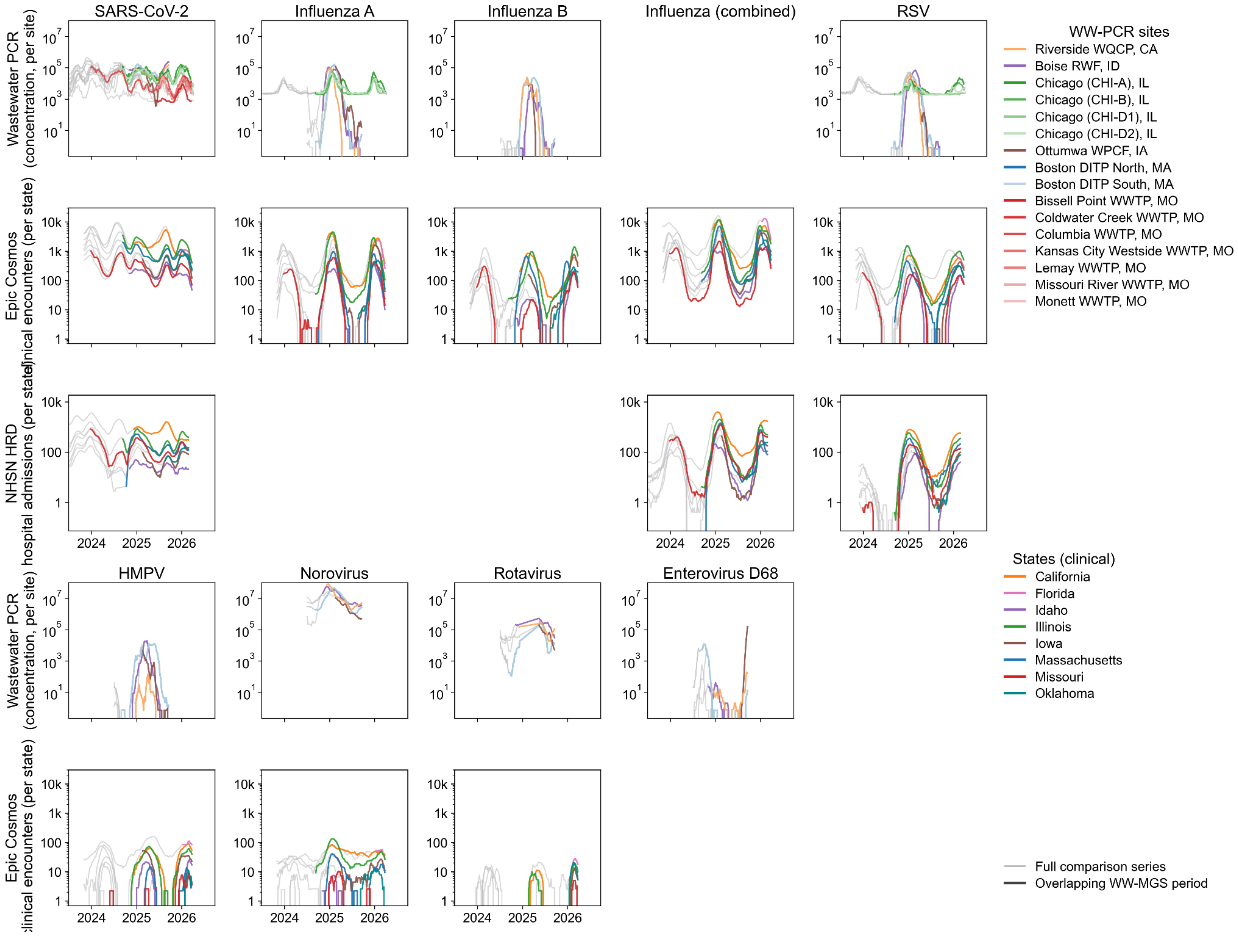


**Supplementary Figure S4. Time series abundance of wastewater PCR (WW-PCR) and clinical data.** Each line is plotted in light grey across its full available history; the portion of the line that overlaps the wastewater metagenomic sequencing (WW-MGS) sampling window for the matched (site, pathogen) or (state, pathogen) is drawn in color. Flat lines in WW-PCR plots indicate lower limits of detection. NHSN, National Healthcare Safety Network.

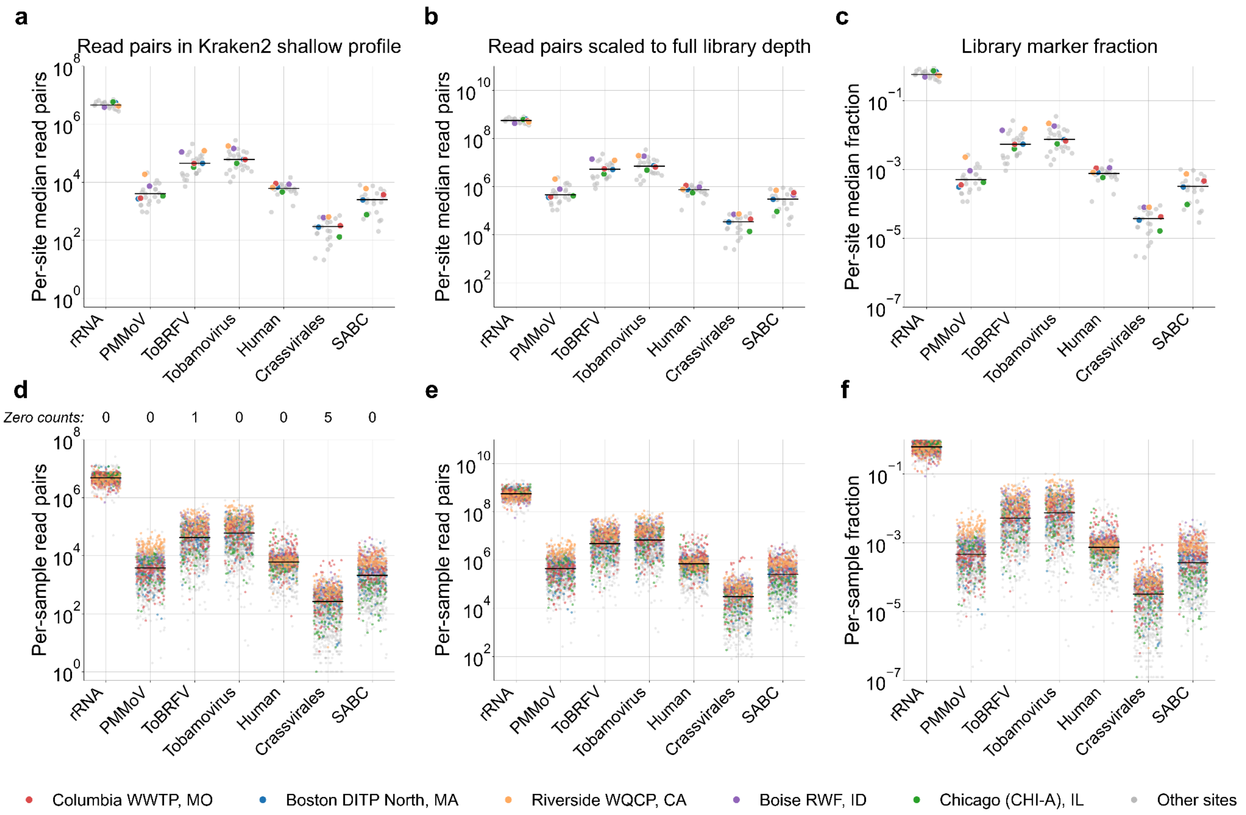


**Supplementary Figure S5. Normalization marker abundance per sample and per site.** Per-marker read counts and library fractions for the seven normalization markers evaluated in this paper. Top row (a–c) — one point per site, taken as the median across that site's samples; bottom row (d–f) — one point per sample. (a, d) Read pairs in the Kraken2 shallow taxonomic profile. (b, e) Read pairs scaled to full library depth (Kraken2 shallow-profile count × total read pairs ÷ profiled read pairs), the denominator used by every marker-based normalization formula in the manuscript. (c, f) Library marker fraction (scaled count ÷ total read pairs), matching main-text Figure 2b. The zero-count tally above panel d reports the number of samples with zero reads at each marker (identical across the three columns). Five sites are highlighted in color (Columbia WWTP, MO; Boston DITP North, MA; Riverside WQCP, CA; Boise RWF, ID; Chicago CHI-A, IL); all other sites are shown in grey. PMMoV, pepper mild mottle virus; ToBRFV, tomato brown rugose fruit virus; SABC, strict-anaerobic-gut-bacteria composite.

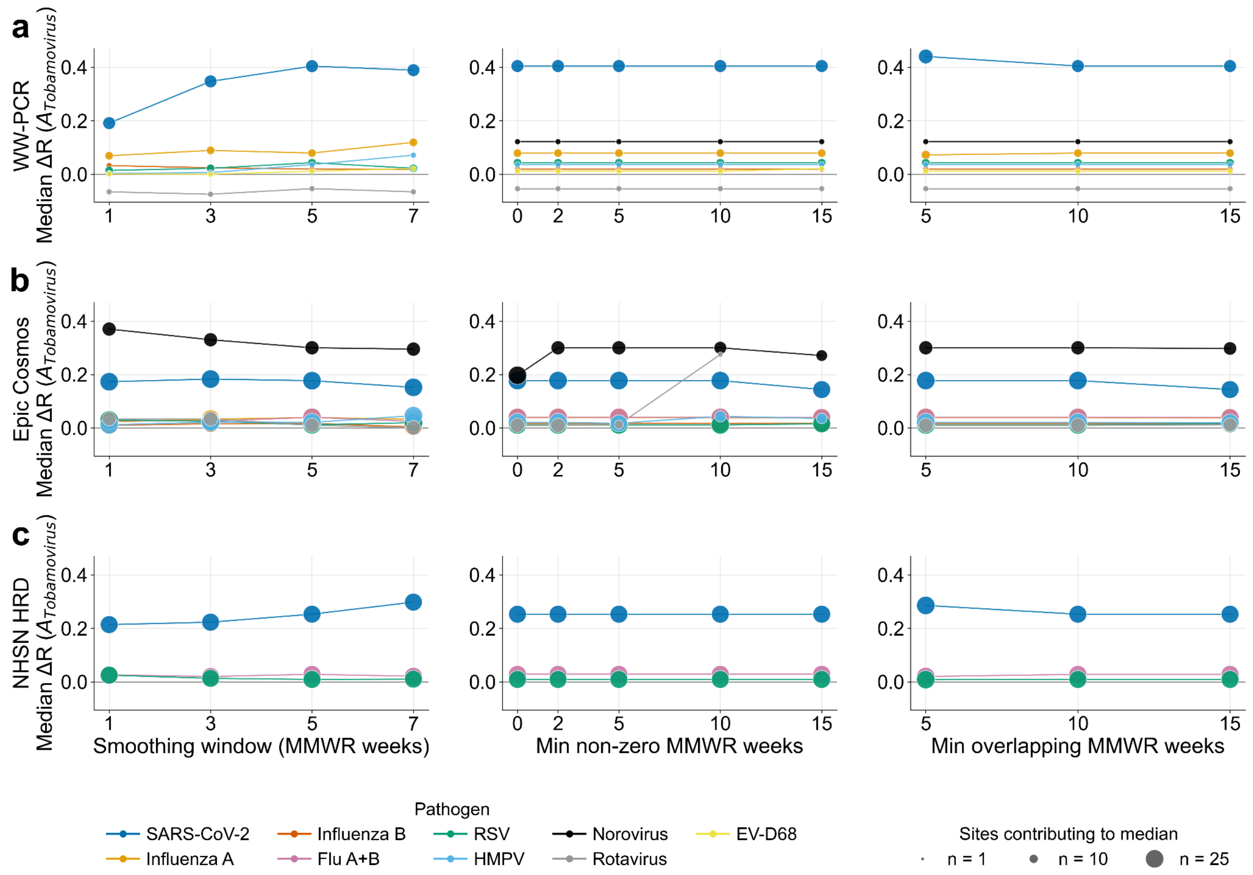


**Supplementary Figure S6. Sensitivity of $A_{Tobamovirus}$ median ΔR to smoothing-window width, non-zero-weeks filter, and minimum-overlap-weeks threshold.** Rows are comparison sources: (a) wastewater PCR (WW-PCR), (b) Epic Cosmos state-level clinical encounters, (c) National Healthcare Safety Network (NHSN) Hospital Respiratory Data (HRD) state-level hospital admissions. Columns are sensitivity sweeps over MMWR smoothing-window width (1, 3, 5, 7 weeks; 1 = raw data; manuscript headlines is 5 weeks); minimum number of non-zero MMWR weeks required on the comparison side (0, 2, 5, 10, 15; the manuscript headline is 2 weeks on each side); minimum total overlapping MMWR weeks between WW-MGS and the comparator (5, 10, 15; the manuscript headline is 10).

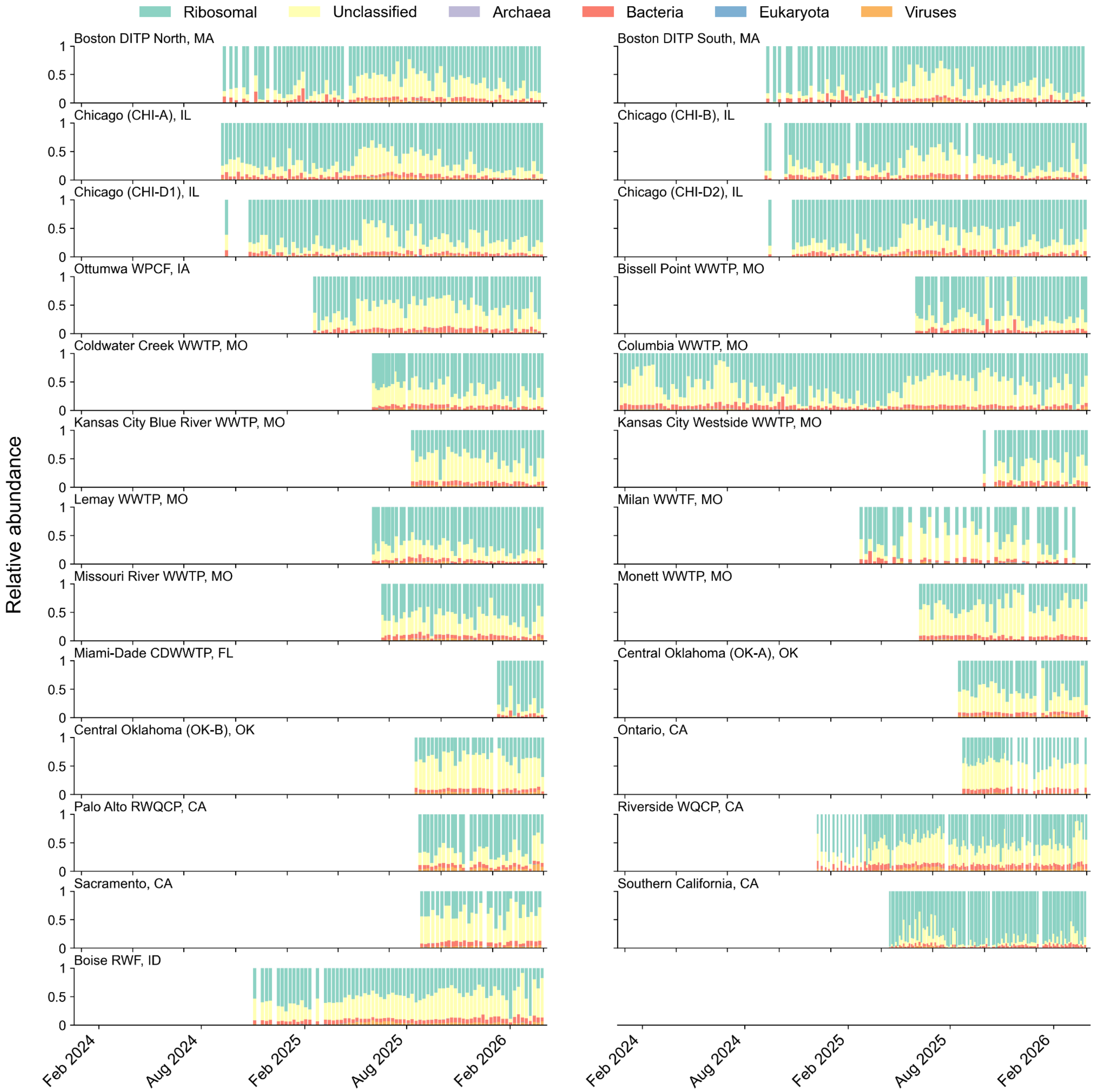


**Supplementary Figure S7. Taxonomic composition over time by site.** Proportion of reads assigned to major taxonomic categories for each sampling location.

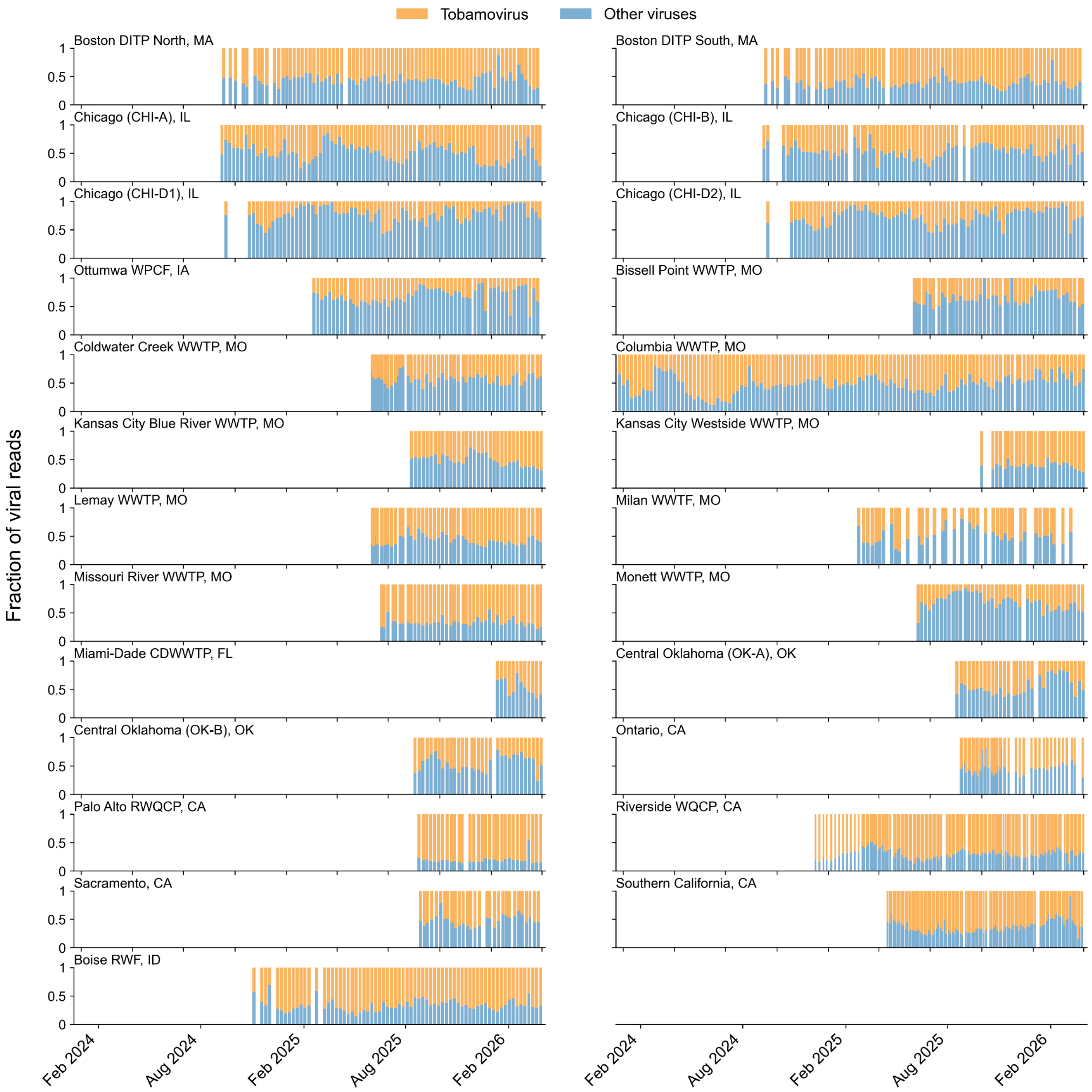


**Supplementary Figure S8. Tobamovirus reads as a fraction of viral reads over time by site.** Fraction of viral reads assigned to the tobamovirus genus for each sampling location as identified by Kraken2 against the Standard database.

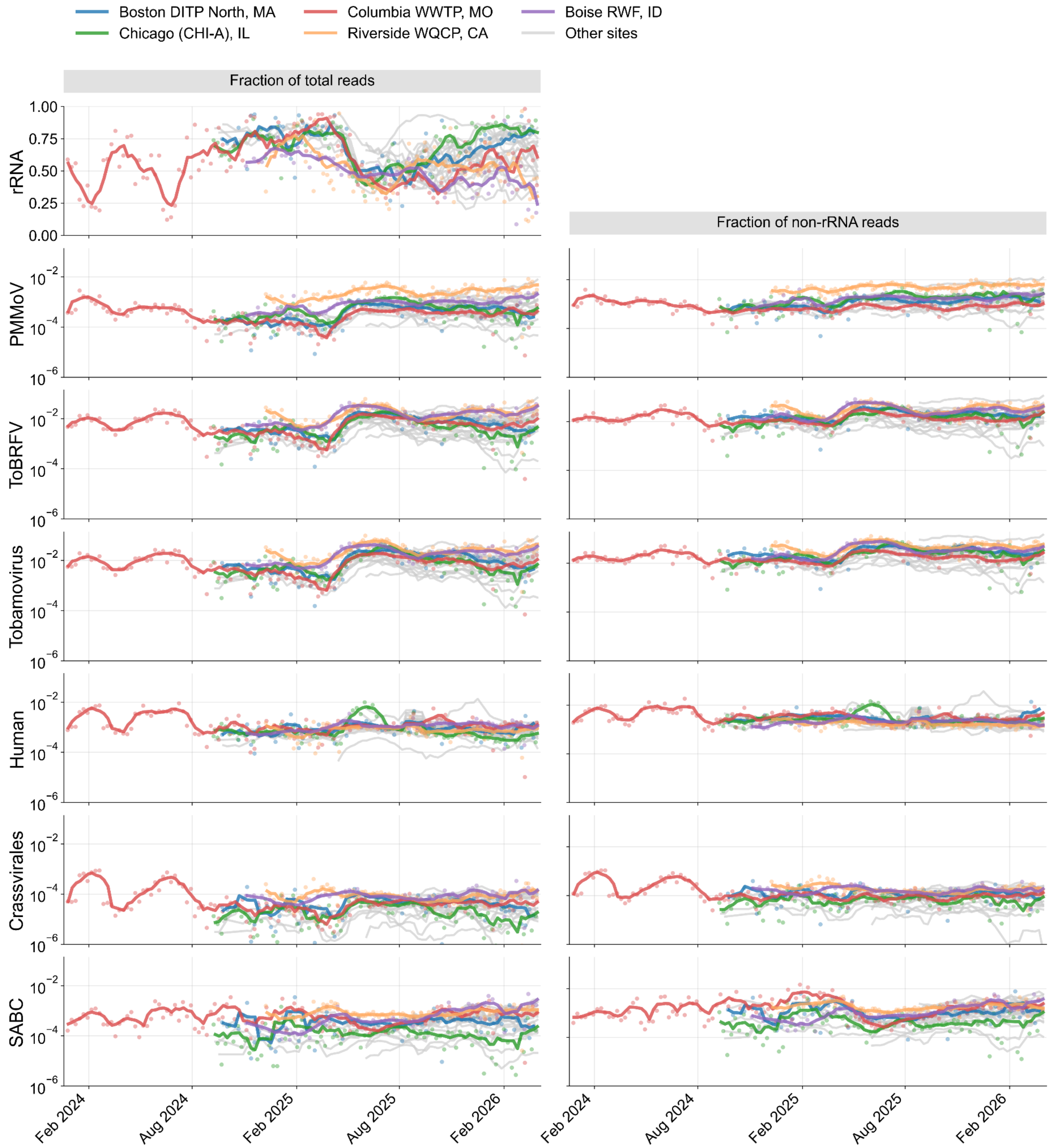


**Supplementary Figure S9. Time series of normalization marker fractions as a fraction of total reads and non-rRNA reads**. Select highlighted sites shown in color; other sites in gray. PMMoV, pepper mild mottle virus; ToBRFV, tomato brown rugose fruit virus; SABC, strict anaerobic gut bacteria composite.

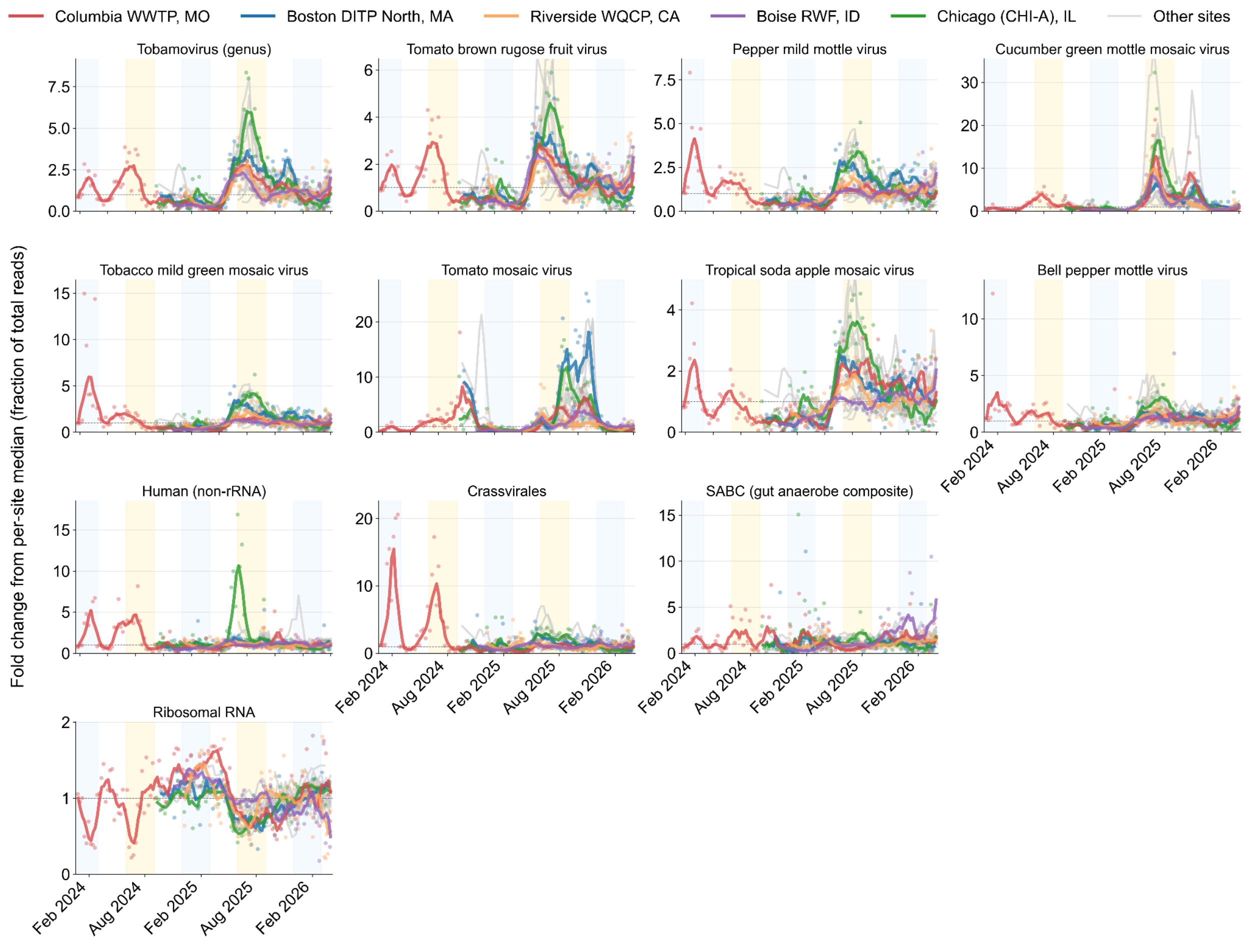


**Supplementary Figure S10. Within-site temporal stability of normalization-marker fractions.** Per-(site, marker) fold change in the marker's fraction of total library read pairs, centred on each site's median fraction across all dates (median computed only on positive values). SABC, strict-anaerobic-gut-bacteria composite.

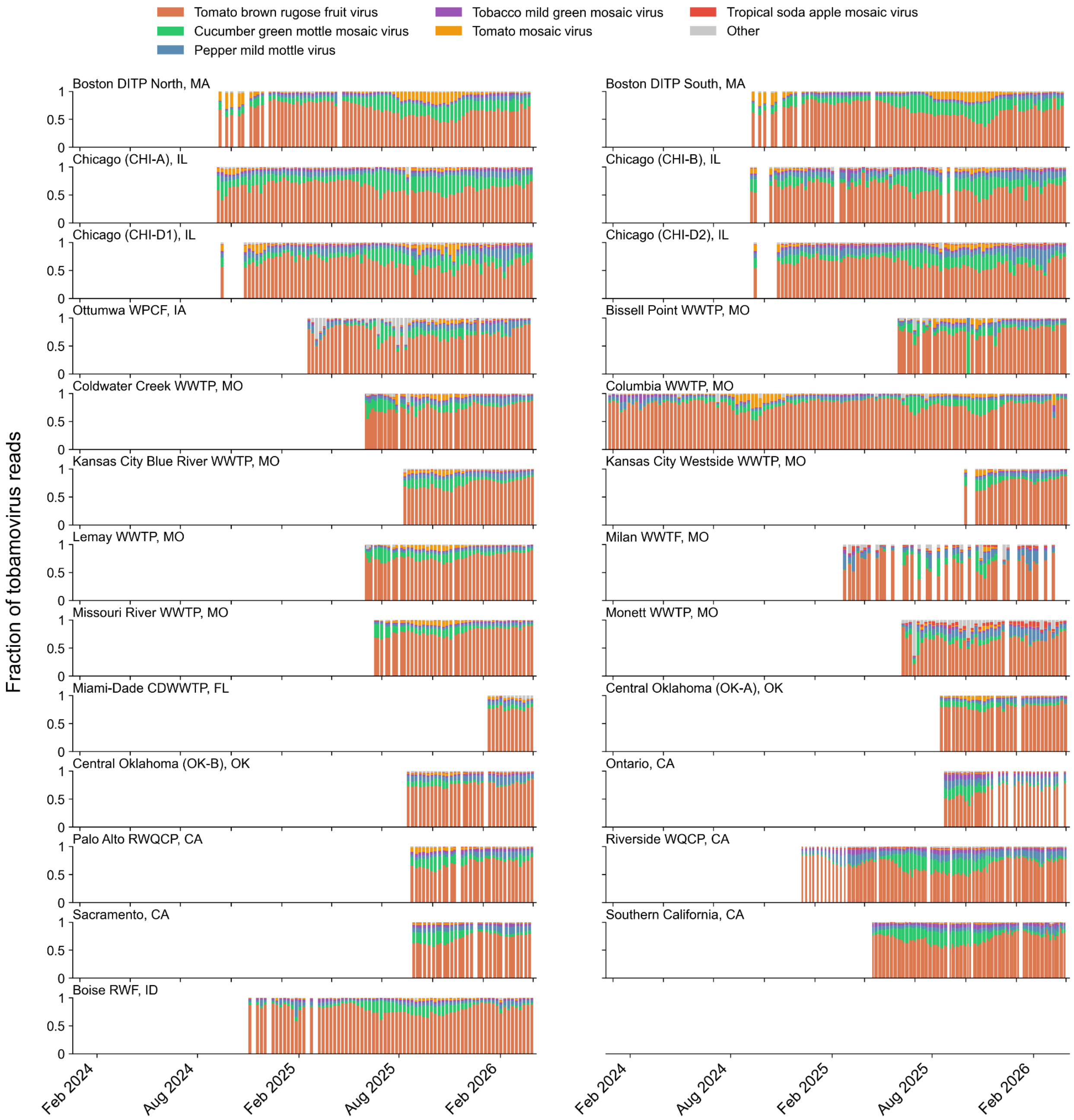


**Supplementary Figure S11. Tobamovirus genus composition over time by site.** Relative abundance of the six most abundant species with tobamovirus genus for each sampling location.

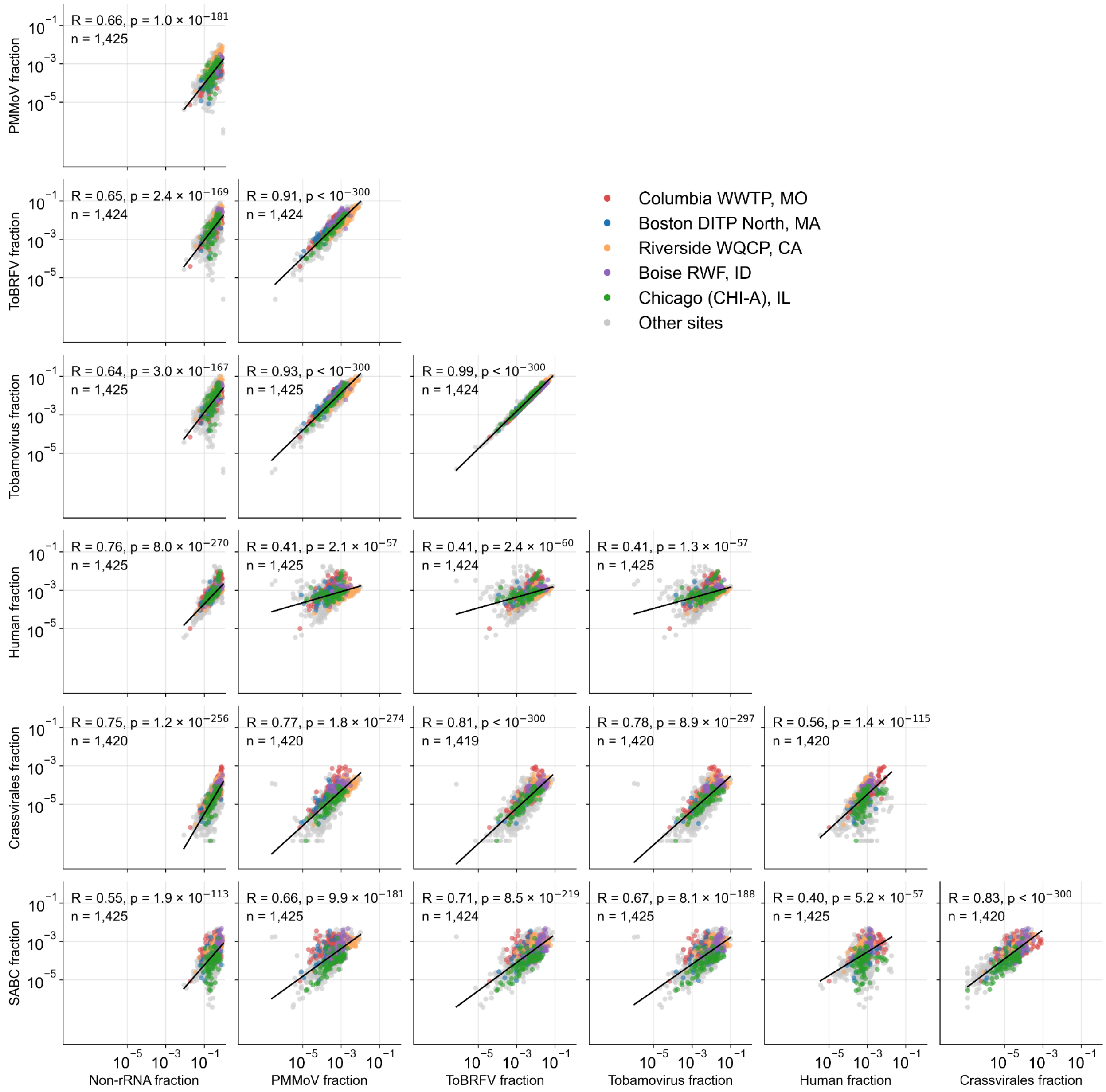


**Supplementary Figure S12. Pairwise correlations between normalization marker fractions across all samples.** Pearson R computed on $\log_{10}$-transformed values. PMMoV, pepper mild mottle virus; ToBRFV, tomato brown rugose fruit virus; SABC, strict anaerobic gut bacteria composite.

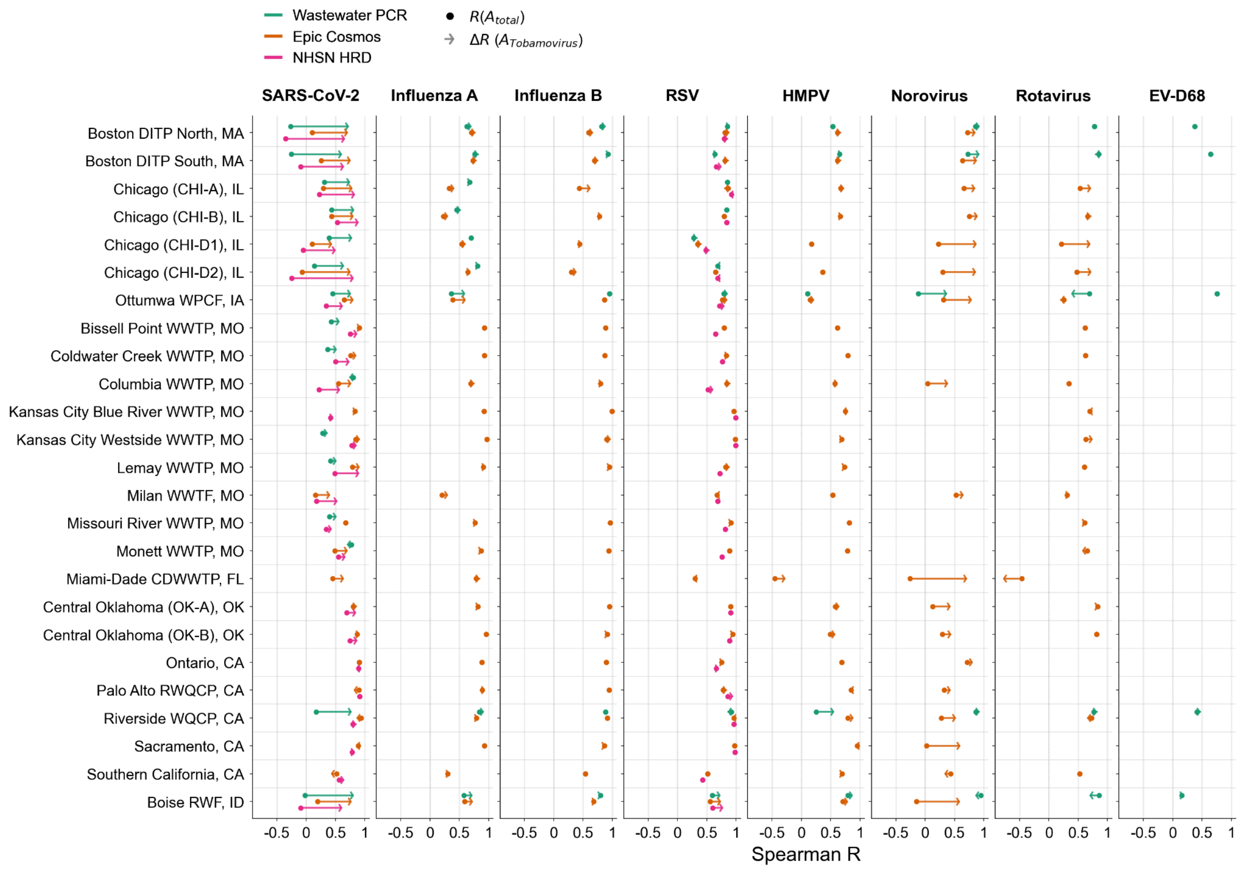


**Supplementary Figure S13. Site-level impact of normalizing wastewater metagenomic sequencing data with tobamovirus genus counts against wastewater PCR and clinical trends across eight pathogens.** Arrows represent change in Spearman R from total read relative abundance ($A_{total}$, baseline) to tobamovirus-normalized abundance ($A_{Tobamovirus}$) for each comparison source (Wastewater PCR, Epic clinical encounters, National Healthcare Safety Network hospital admissions).

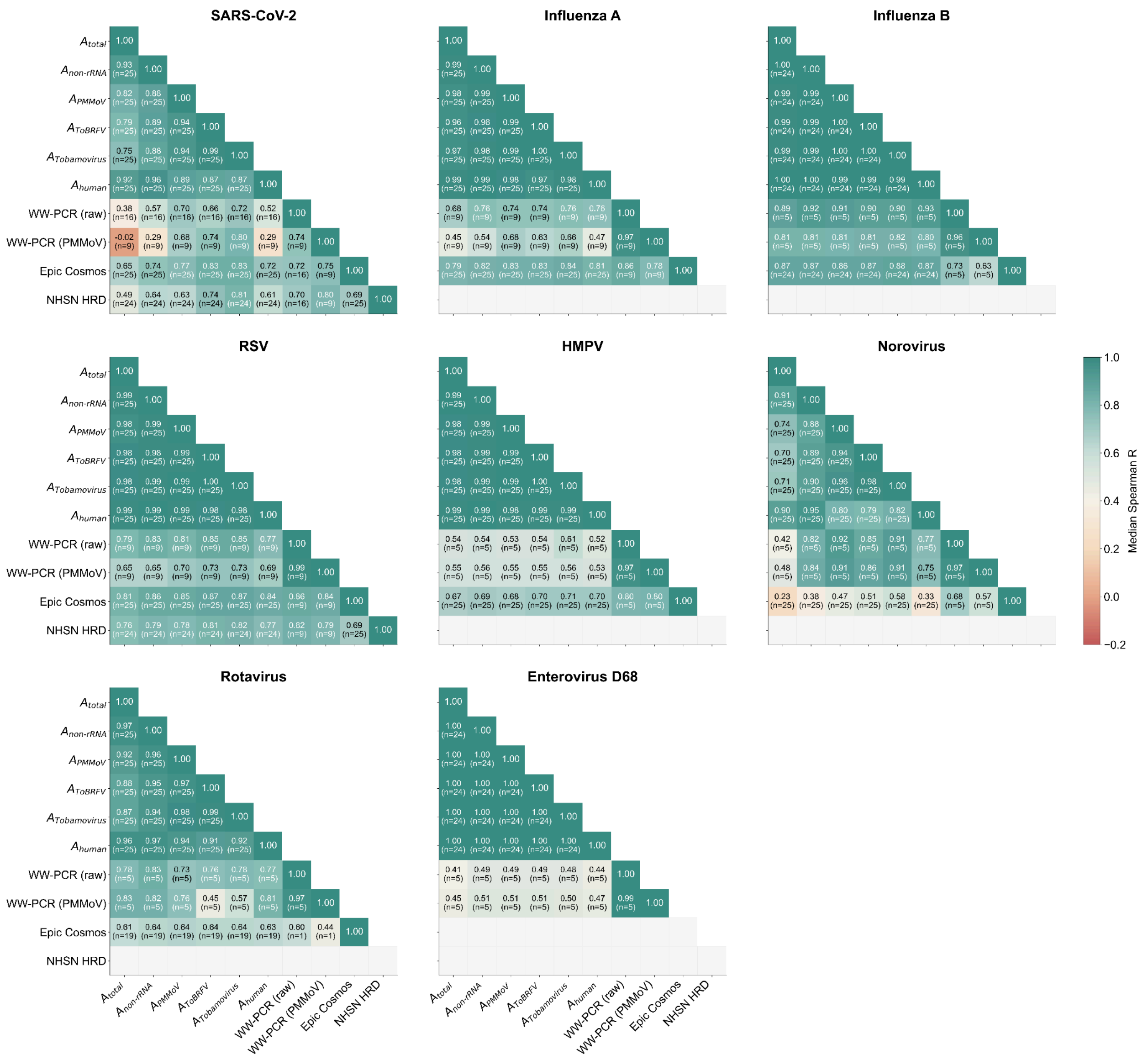

**Supplementary Figure S14. Cross-source agreement between wastewater metagenomic sequencing, wastewater PCR, and clinical comparators.** Per-pathogen heatmaps of pairwise median per-site Spearman R between WW-MGS signals, wastewater PCR (raw and PMMoV-normalized concentrations), Epic Cosmos clinical encounters, and NHSN Hospital Respiratory Data (HRD) admissions. Each cell shows the median Spearman R across all sites where both signals are available, with the number of contributing sites (n) in parentheses. Wastewater PCR cells are restricted to the WastewaterSCAN sites and NWSS sites used in the main WW-MGS PCR analysis (Supplementary Table S1). Gray cells indicate combinations with no data. PMMoV, pepper mild mottle virus; ToBRFV, tomato brown rugose fruit virus; NHSN HRD, National Healthcare Safety Network Hospital Respiratory Data; WW-PCR, wastewater PCR; WW-MGS, wastewater metagenomic sequencing.

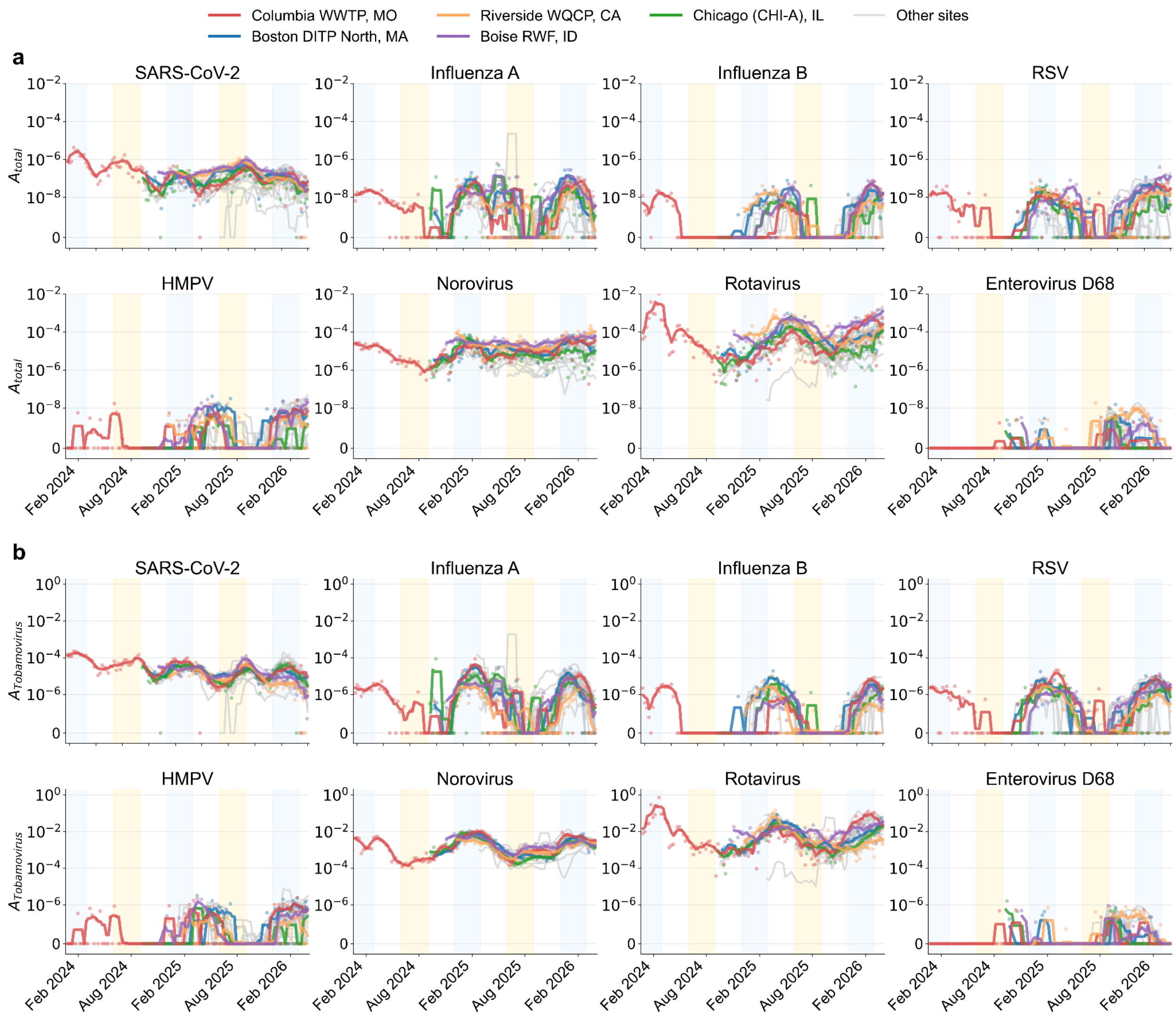


**Supplementary Figure S15. Per site pathogen abundance trends in wastewater metagenomic sequencing (WW-MGS) data.** (a) Total read relative abundance ($A_{total}$) of pathogens in WW-MGS data. (b) Tobamovirus-normalized abundance ($A_{Tobamovirus}$) of pathogens in WW-MGS data. Select highlighted sites shown in color; other sites in gray.

**Supplementary Table S1. Wastewater PCR (WW-PCR) data sources mapped to CASPER wastewater metagenomic sequencing (WW-MGS) sites.** For each of the 25 CASPER sampling sites with WW-MGS coverage, columns report the matched National Wastewater Surveillance System (NWSS) sewershed identifier (where one exists and is permissible to share publically), the wastewater PCR source used in the analysis, the WastewaterSCAN sewershed name where applicable, the set of pathogen targets quantified by that source at the site, and the WW-PCR normalization variants reported alongside the raw concentration (PMMoV-normalized concentration for WastewaterSCAN; PMMoV-normalized and flow-population-normalized concentrations for NWSS). Where both WastewaterSCAN and NWSS coverage existed, WastewaterSCAN was used to avoid double-counting. Em-dashes indicate no usable WW-PCR data at the corresponding site or comparator after applying minimum-overlap criteria (see Methods); 9 of the 25 sites had no usable WW-PCR comparison data. IAV (influenza A virus); IBV (influenza B virus); PMMoV, pepper mild mottle virus. NWSS sewershed identifiers are CDC NWSS internal sewershed IDs used by the Socrata API (e.g., (CDC, 2025c)).

| Site | NWSS sewershed ID | WW-PCR source | WastewaterSCAN site | Pathogen targets | WW-PCR normalization targets |
|---|---|---|---|---|---|
| Boston DITP North, MA | 742 | WastewaterSCAN | Deer Island Treatment Plant (Boston, MA) | SARS-CoV-2, IAV, IBV, RSV, HMPV, Rotavirus, EV-D68, Norovirus GII | PMMoV |
| Boston DITP South, MA | 742 | WastewaterSCAN | Deer Island Treatment Plant (Boston, MA) | SARS-CoV-2, IAV, IBV, RSV, HMPV, Rotavirus, EV-D68, Norovirus GII | PMMoV |
| Chicago (CHI-A), IL | 419 | NWSS state/territory | — | SARS-CoV-2, IAV, RSV | PMMoV, Flow-population |
| Chicago (CHI-B), IL | 413 | NWSS state/territory | — | SARS-CoV-2, IAV, RSV | PMMoV, Flow-population |
| Chicago (CHI-D1), IL | 423 | NWSS state/territory | — | SARS-CoV-2, IAV, RSV | PMMoV, Flow-population |
| Chicago (CHI-D2), IL | 429 | NWSS state/territory | — | SARS-CoV-2, IAV, RSV | PMMoV, Flow-population |
| Ottumwa WPCF, IA | 541 | WastewaterSCAN | Ottumwa WPCF (Ottumwa, IA) | SARS-CoV-2, IAV, IBV, RSV, HMPV, Rotavirus, EV-D68, Norovirus GII | PMMoV |
| Bissell Point WWTP, MO | 1139 | NWSS state/territory | — | SARS-CoV-2 | Flow-population |
| Coldwater Creek WWTP, MO | 1123 | NWSS state/territory | — | SARS-CoV-2 | Flow-population |
| Columbia WWTP, MO | 1045 | NWSS state/territory | — | SARS-CoV-2, IAV, RSV | Flow-population |
| Kansas City Blue River WWTP, MO | 1074 | — | — | — | — |
| Kansas City Westside WWTP, MO | 1075 | NWSS state/territory | — | SARS-CoV-2, IAV, RSV | Flow-population |
| Lemay WWTP, MO | | | | | |
| Milan WWTF, MO | — | — | — | — | — |
| Missouri River WWTP, MO | 1128 | NWSS state/territory | — | SARS-CoV-2 | Flow-population |
| Monett WWTP, MO | 1041 | NWSS state/territory | — | SARS-CoV-2, IAV, RSV | Flow-population |
| Miami-Dade CDWWTP, FL | 310 | — | — | — | — |
| Central Oklahoma (OK-A), OK | 1704 | — | — | — | — |
| Central Oklahoma (OK-B), OK | 1706 | — | — | — | — |
| Ontario, CA | 143 | — | — | — | — |
| Palo Alto RWQCP, CA | 175 | — | — | — | — |
| Riverside WQCP, CA | 138 | WastewaterSCAN | Riverside Water Quality Control Plant (Riverside, CA) | SARS-CoV-2, IAV, IBV, RSV, HMPV, Rotavirus, EV-D68, Norovirus GII | PMMoV |
| Sacramento, CA | 140 | — | — | — | — |
| Southern California, CA | — | — | — | — | — |
| Boise RWF, ID | 372 | WastewaterSCAN | West Boise Water Renewal Facility; Lander Street Water Renewal Facility (Boise, ID) | SARS-CoV-2, IAV, IBV, RSV, HMPV, Rotavirus, EV-D68, Norovirus GII | PMMoV |

**Supplementary Table S2. Taxonomic and case-definition matching between wastewater metagenomic sequencing (WW-MGS) and each comparison source.** For each pathogen (rows), the WW-MGS taxonomic identifier (NCBI taxid; reads assigned to this taxid or any of its descendants are counted toward the pathogen) is paired with the corresponding target used by each external comparator (columns): the PCR amplification target gene for WastewaterSCAN ddRT-PCR on wastewater solids (Boehm et al., 2026) and state/territory National Wastewater Surveillance System (NWSS) qPCR / ddPCR on wastewater influent (CDC, 2025a, 2025b, 2025c); the ICD-10 diagnosis code or lab-confirmed test result used to define a case in Epic Cosmos clinical encounters (Tarabichi et al., 2021); and the admission category reported in the CDC National Healthcare Safety Network (NHSN) Hospital Respiratory Data (HRD) system (CDC, 2024). Influenza A and B are reported separately in some sources but combined in others, so we list a separate "Influenza (combined)" row that pairs the WW-MGS A+B-summed taxid pair (197911 + 197912) against the comparators that report combined flu (Cosmos' ICD J09–J11 group and NHSN HRD's combined flu admissions). Em-dashes indicate pathogen × source combinations with no available comparator data within the CASPER sampling window.

| Pathogen | WW-MGS taxid <> WastewaterSCAN | WW-MGS taxid <> NWSS | WW-MGS taxid <> Epic Cosmos | WW-MGS taxid <> NHSN HRD |
|---|---|---|---|---|
| SARS-CoV-2 | 2697049 <> N gene | 2697049 <> N gene | 2697049 <> ICD U07 | 2697049 <> COVID-19 admissions |
| Influenza A | 197911<> M gene | 197911 <> M gene | 197911 <> Lab-confirmed Flu A | — |
| Influenza B | 197912 <> L gene | — | 197912 <> Lab-confirmed Flu B | — |
| Influenza (combined) | — | — | 197911 + 197912 <> ICD J09–J11 | 197911 + 197912 <> Flu admissions |
| RSV | 3049954 <> N gene | 3049954 <> RSV A & B | 3049954 <> ICD B97.4, J12.1, J20.5, J21.0 | 3049954 <> RSV admissions |
| HMPV | 3048148 <> L gene | — | 3048148 <> ICD B97.81, J12.3, J21.1 | — |
| Norovirus | 122929 <> ORF1–2 junction | — | 11983 <> ICD A08.11 | — |
| Rotavirus | 28875 <> NSP3 | — | 28875 <> ICD A08.0 | — |
| Enterovirus D68 | 42789 <> VP1 | — | — | — |

**Supplementary Table S3. Zero denominator counts for each normalization approach.**

| Normalization | n samples | n dropped |
|---|---|---|
| non-rRNA | 1,425 | 0 |
| PMMoV | 1,425 | 0 |
| ToBRFV | 1,425 | 1 |
| Tobamovirus | 1,425 | 0 |
| Human | 1,425 | 0 |
| Crassvirales | 1,425 | 5 |
| SABC | 1,425 | 0 |

**Supplementary Table S4. Normalization performance across comparison sources by pathogen.** For each pathogen × normalization method (rows), columns report site-level Spearman R between the wastewater metagenomic sequencing (WW-MGS) abundance estimate and three reference sources (wastewater PCR (WW-PCR), Epic Cosmos clinical encounters, National Healthcare Safety Network (NHSN) Hospital Respiratory Data (HRD)), summarized as the mean and median R across sites, the fraction of sites where the normalization improved over the $A_{total}$ baseline and the mean and median per-site change in R relative to baseline (ΔR). P values are from a two-sided Wilcoxon signed-rank test against the null of median ΔR = 0; the test runs on the per-site ΔR distribution for WW-PCR cells (where each site has its own paired comparison series) and on per-state mean ΔR for Epic Cosmos and NHSN cells (where sites within a state share one state-aggregated comparison series and ΔR values are clustered), and p-values are Benjamini–Hochberg corrected within each (pathogen × source) combination across normalization methods. Cells with fewer than six effective units have a blank P value. Empty cells indicate pathogen–source combinations with no overlapping data.

| | Wastewater PCR | | | | | | Epic Cosmos clinical encounters | | | | | | NHSN HRD hospital admissions | | | | | |
|---|---|---|---|---|---|---|---|---|---|---|---|---|---|---|---|---|---|---|
| | Mean R | Med. R | Improved | Mean ΔR | Med. ΔR | p | Mean R | Med. R | Improved | Mean ΔR | Med. ΔR | p | Mean R | Med. R | Improved | Mean ΔR | Med. ΔR | p |
| **SARS-CoV-2** | | | | | | | | | | | | | | | | | | |
| $A_{total}$ | 0.302 | 0.380 | — | — | — | — | 0.570 | 0.652 | — | — | — | — | 0.412 | 0.495 | — | — | — | — |
| $A_{non\text{-}rRNA}$ | 0.508 | 0.574 | 13/16 | 0.206 | 0.198 | 1.02E-03 | 0.682 | 0.738 | 19/25 | 0.112 | 0.048 | 9.57E-02 | 0.584 | 0.638 | 22/24 | 0.171 | 0.143 | 2.73E-02 |
| $A_{PMMoV}$ | 0.626 | 0.703 | 12/16 | 0.324 | 0.352 | 4.7E-03 | 0.741 | 0.767 | 17/25 | 0.171 | 0.100 | 5.47E-02 | 0.649 | 0.626 | 19/24 | 0.237 | 0.156 | 4.37E-02 |
| $A_{ToBRFV}$ | 0.671 | 0.663 | 14/16 | 0.369 | 0.401 | 1.02E-03 | 0.791 | 0.832 | 19/25 | 0.221 | 0.173 | 5.47E-02 | 0.736 | 0.742 | 20/24 | 0.324 | 0.213 | 2.73E-02 |
| $A_{Tobamovirus}$ | 0.706 | 0.719 | 14/16 | 0.404 | 0.404 | 1.02E-03 | 0.793 | 0.831 | 20/25 | 0.223 | 0.177 | 5.47E-02 | 0.769 | 0.812 | 22/24 | 0.357 | 0.253 | 2.73E-02 |
| $A_{human}$ | 0.456 | 0.517 | 14/16 | 0.154 | 0.120 | 1.02E-03 | 0.667 | 0.718 | 20/25 | 0.097 | 0.042 | 1.09E-01 | 0.543 | 0.611 | 19/24 | 0.131 | 0.129 | 2.73E-02 |
| $A_{Crassvirales}$ | 0.513 | 0.575 | 12/16 | 0.211 | 0.209 | 8.9E-03 | 0.688 | 0.694 | 17/25 | 0.118 | 0.076 | 1.28E-01 | 0.578 | 0.629 | 16/24 | 0.166 | 0.104 | 2.55E-01 |
| $A_{SABC}$ | 0.441 | 0.499 | 11/16 | 0.139 | 0.176 | 2.14E-02 | 0.593 | 0.583 | 14/25 | 0.023 | 0.037 | 4.61E-01 | 0.461 | 0.545 | 13/24 | 0.049 | 0.003 | 4.69E-01 |
| **Influenza A** | | | | | | | | | | | | | | | | | | |
| $A_{total}$ | 0.648 | 0.676 | — | — | — | — | 0.710 | 0.791 | — | — | — | — | | | | | | |
| $A_{non\text{-}rRNA}$ | 0.723 | 0.763 | 9/9 | 0.075 | 0.058 | 1.37E-02 | 0.745 | 0.817 | 22/25 | 0.035 | 0.022 | 2.19E-02 | | | | | | |
| $A_{PMMoV}$ | 0.737 | 0.740 | 8/9 | 0.089 | 0.056 | 2.73E-02 | 0.753 | 0.827 | 22/25 | 0.043 | 0.038 | 1.37E-02 | | | | | | |
| $A_{ToBRFV}$ | 0.742 | 0.740 | 8/9 | 0.094 | 0.068 | 1.37E-02 | 0.768 | 0.830 | 23/25 | 0.058 | 0.042 | 1.37E-02 | | | | | | |
| $A_{Tobamovirus}$ | 0.752 | 0.764 | 8/9 | 0.103 | 0.079 | 1.37E-02 | 0.772 | 0.842 | 23/25 | 0.062 | 0.039 | 1.37E-02 | | | | | | |
| $A_{human}$ | 0.702 | 0.759 | 9/9 | 0.054 | 0.028 | 1.37E-02 | 0.728 | 0.809 | 21/25 | 0.018 | 0.019 | 4.56E-02 | | | | | | |
| $A_{Crassvirales}$ | 0.692 | 0.723 | 7/9 | 0.044 | 0.074 | 1.91E-01 | 0.745 | 0.801 | 19/25 | 0.035 | 0.029 | 1.37E-02 | | | | | | |
| $A_{SABC}$ | 0.662 | 0.667 | 5/9 | 0.014 | 0.013 | 9.1E-01 | 0.691 | 0.754 | 13/25 | -0.019 | 0.000 | 9.38E-01 | | | | | | |
| **Influenza B** | | | | | | | | | | | | | | | | | | |
| $A_{total}$ | 0.876 | 0.887 | — | — | — | — | 0.788 | 0.869 | — | — | — | — | | | | | | |
| $A_{non\text{-}rRNA}$ | 0.895 | 0.924 | 3/5 | 0.020 | 0.013 | — | 0.810 | 0.870 | 16/23 | 0.021 | 0.004 | 1.09E-01 | | | | | | |
| $A_{PMMoV}$ | 0.899 | 0.911 | 3/5 | 0.023 | 0.024 | — | 0.812 | 0.867 | 10/23 | 0.023 | 0.000 | 8.12E-01 | | | | | | |
| $A_{ToBRFV}$ | 0.900 | 0.898 | 4/5 | 0.024 | 0.017 | — | 0.824 | 0.876 | 17/23 | 0.036 | 0.013 | 1.09E-01 | | | | | | |
| $A_{Tobamovirus}$ | 0.901 | 0.899 | 4/5 | 0.025 | 0.019 | — | 0.825 | 0.882 | 17/23 | 0.037 | 0.017 | 1.09E-01 | | | | | | |
| $A_{human}$ | 0.880 | 0.926 | 2/5 | 0.004 | 0.000 | — | 0.804 | 0.871 | 15/23 | 0.016 | 0.003 | 2.73E-01 | | | | | | |
| $A_{Crassvirales}$ | 0.898 | 0.930 | 3/5 | 0.023 | 0.025 | — | 0.807 | 0.870 | 10/23 | 0.019 | -0.002 | 8.02E-01 | | | | | | |
| $A_{SABC}$ | 0.884 | 0.928 | 3/5 | 0.008 | 0.000 | — | 0.790 | 0.876 | 11/23 | 0.002 | 0.000 | 8.02E-01 | | | | | | |
| **Influenza (combined)** | | | | | | | | | | | | | | | | | | |
| $A_{total}$ | | | | | | | 0.722 | 0.800 | — | — | — | — | 0.724 | 0.797 | — | — | — | — |
| $A_{non\text{-}rRNA}$ | | | | | | | 0.762 | 0.852 | 20/25 | 0.040 | 0.021 | 4.37E-02 | 0.766 | 0.814 | 17/24 | 0.042 | 0.018 | 5.47E-02 |
| $A_{PMMoV}$ | | | | | | | 0.760 | 0.830 | 20/25 | 0.038 | 0.021 | 2.73E-02 | 0.776 | 0.832 | 20/24 | 0.052 | 0.027 | 5.47E-02 |
| $A_{ToBRFV}$ | | | | | | | 0.783 | 0.841 | 20/25 | 0.062 | 0.045 | 2.73E-02 | 0.776 | 0.825 | 18/24 | 0.052 | 0.025 | 5.47E-02 |
| $A_{Tobamovirus}$ | | | | | | | 0.787 | 0.851 | 19/25 | 0.065 | 0.040 | 2.73E-02 | 0.781 | 0.836 | 17/24 | 0.057 | 0.028 | 5.47E-02 |
| $A_{human}$ | | | | | | | 0.743 | 0.834 | 19/25 | 0.021 | 0.018 | 4.56E-02 | 0.750 | 0.811 | 19/24 | 0.026 | 0.014 | 5.47E-02 |
| $A_{Crassvirales}$ | | | | | | | 0.761 | 0.855 | 19/25 | 0.039 | 0.011 | 2.73E-02 | 0.761 | 0.825 | 16/24 | 0.037 | 0.011 | 5.47E-02 |
| $A_{SABC}$ | | | | | | | 0.704 | 0.810 | 12/25 | -0.017 | 0.000 | 2.97E-01 | 0.718 | 0.778 | 11/24 | -0.006 | -0.003 | 3.75E-01 |
| **RSV** | | | | | | | | | | | | | | | | | | |
| $A_{total}$ | 0.716 | 0.789 | — | — | — | — | 0.777 | 0.811 | — | — | — | — | 0.762 | 0.763 | — | — | — | — |
| $A_{non\text{-}rRNA}$ | 0.744 | 0.833 | 6/9 | 0.029 | 0.038 | 2.3E-01 | 0.787 | 0.855 | 17/25 | 0.010 | 0.013 | 2.73E-01 | 0.785 | 0.790 | 13/24 | 0.023 | 0.013 | 2.73E-02 |
| $A_{PMMoV}$ | 0.745 | 0.812 | 5/9 | 0.030 | 0.036 | 2.3E-01 | 0.791 | 0.853 | 14/25 | 0.014 | 0.004 | 1.82E-01 | 0.790 | 0.780 | 17/24 | 0.029 | 0.004 | 2.73E-02 |
| $A_{ToBRFV}$ | 0.756 | 0.850 | 7/9 | 0.040 | 0.012 | 2.37E-01 | 0.808 | 0.869 | 17/25 | 0.031 | 0.011 | 1.82E-01 | 0.797 | 0.811 | 17/24 | 0.035 | 0.008 | 2.73E-02 |
| $A_{Tobamovirus}$ | 0.759 | 0.851 | 7/9 | 0.043 | 0.043 | 2.3E-01 | 0.809 | 0.868 | 18/25 | 0.032 | 0.012 | 1.82E-01 | 0.800 | 0.816 | 18/24 | 0.039 | 0.009 | 2.73E-02 |
| $A_{human}$ | 0.735 | 0.770 | 4/9 | 0.019 | -0.010 | 4.26E-01 | 0.771 | 0.843 | 15/25 | -0.006 | 0.005 | 5.38E-01 | 0.783 | 0.775 | 14/24 | 0.021 | 0.010 | 5.47E-02 |
| $A_{Crassvirales}$ | 0.743 | 0.809 | 6/9 | 0.028 | 0.051 | 2.3E-01 | 0.791 | 0.847 | 16/25 | 0.014 | 0.012 | 2.73E-01 | 0.790 | 0.794 | 15/24 | 0.029 | 0.003 | 4.37E-02 |
| $A_{SABC}$ | 0.743 | 0.830 | 6/9 | 0.028 | 0.019 | 2.3E-01 | 0.758 | 0.816 | 13/25 | -0.019 | 0.008 | 6.41E-01 | 0.773 | 0.805 | 11/24 | 0.011 | -0.003 | 3.75E-01 |
| **HMPV** | | | | | | | | | | | | | | | | | | |
| $A_{total}$ | 0.470 | 0.538 | — | — | — | 0.595 | 0.669 | — | — | — | 0.595 | — | | | | | | |
| $A_{non\text{-}rRNA}$ | 0.507 | 0.544 | 4/5 | 0.038 | 0.028 | 0.619 | 0.691 | 16/25 | 0.024 | 0.005 | 0.619 | 2.73E-02 | | | | | | |
| $A_{PMMoV}$ | 0.528 | 0.535 | 3/5 | 0.058 | 0.038 | 0.615 | 0.678 | 19/25 | 0.021 | 0.012 | 0.615 | 3.28E-02 | | | | | | |
| $A_{ToBRFV}$ | 0.551 | 0.540 | 4/5 | 0.081 | 0.032 | 0.632 | 0.696 | 17/25 | 0.037 | 0.018 | 0.632 | 2.73E-02 | | | | | | |
| $A_{Tobamovirus}$ | 0.568 | 0.614 | 4/5 | 0.099 | 0.036 | 0.637 | 0.705 | 17/25 | 0.042 | 0.021 | 0.637 | 2.73E-02 | | | | | | |
| $A_{human}$ | 0.472 | 0.517 | 2/5 | 0.002 | -0.006 | 0.617 | 0.699 | 15/25 | 0.023 | 0.006 | 0.617 | 2.73E-02 | | | | | | |
| $A_{Crassvirales}$ | 0.497 | 0.540 | 4/5 | 0.027 | 0.031 | 0.615 | 0.694 | 18/25 | 0.020 | 0.016 | 0.615 | 6.38E-02 | | | | | | |
| $A_{SABC}$ | 0.471 | 0.528 | 2/5 | 0.001 | -0.010 | 0.599 | 0.670 | 14/25 | 0.005 | 0.002 | 0.599 | 5.47E-01 | | | | | | |
| **Norovirus** | | | | | | | | | | | | | | | | | | |
| $A_{total}$ | 0.658 | 0.861 | — | — | — | — | 0.333 | 0.304 | — | — | — | — | | | | | | |
| $A_{non\text{-}rRNA}$ | 0.898 | 0.934 | 5/5 | 0.240 | 0.105 | — | 0.483 | 0.470 | 16/18 | 0.150 | 0.153 | 1.73E-01 | | | | | | |
| $A_{PMMoV}$ | 0.956 | 0.974 | 4/5 | 0.298 | 0.129 | — | 0.573 | 0.601 | 14/18 | 0.240 | 0.213 | 5.47E-02 | | | | | | |
| $A_{ToBRFV}$ | 0.816 | 0.944 | 4/5 | 0.158 | 0.120 | — | 0.694 | 0.740 | 17/18 | 0.361 | 0.284 | 2.73E-02 | | | | | | |
| $A_{Tobamovirus}$ | 0.845 | 0.979 | 4/5 | 0.187 | 0.122 | — | 0.712 | 0.742 | 17/18 | 0.379 | 0.300 | 2.73E-02 | | | | | | |
| $A_{human}$ | 0.763 | 0.924 | 4/5 | 0.105 | 0.081 | — | 0.465 | 0.451 | 16/18 | 0.132 | 0.128 | 1.73E-01 | | | | | | |
| $A_{Crassvirales}$ | 0.764 | 0.907 | 4/5 | 0.106 | 0.103 | — | 0.471 | 0.444 | 13/18 | 0.138 | 0.140 | 6.84E-02 | | | | | | |
| $A_{SABC}$ | 0.501 | 0.625 | 0/5 | -0.157 | -0.106 | — | 0.383 | 0.496 | 11/18 | 0.050 | 0.081 | 9.45E-01 | | | | | | |
| **Rotavirus** | | | | | | | | | | | | | | | | | | |
| $A_{total}$ | 0.789 | 0.775 | — | — | — | — | 0.508 | 0.610 | — | — | — | — | | | | | | |
| $A_{non\text{-}rRNA}$ | 0.805 | 0.833 | 3/5 | 0.016 | 0.012 | — | 0.574 | 0.639 | 14/19 | 0.065 | 0.014 | 2.19E-01 | | | | | | |
| $A_{PMMoV}$ | 0.798 | 0.731 | 3/5 | 0.009 | 0.005 | — | 0.551 | 0.644 | 13/19 | 0.043 | 0.014 | 1E+00 | | | | | | |
| $A_{ToBRFV}$ | 0.662 | 0.759 | 1/5 | -0.127 | -0.093 | — | 0.559 | 0.640 | 13/19 | 0.051 | 0.010 | 1E+00 | | | | | | |
| $A_{Tobamovirus}$ | 0.683 | 0.782 | 2/5 | -0.106 | -0.054 | — | 0.554 | 0.641 | 12/19 | 0.045 | 0.010 | 1E+00 | | | | | | |
| $A_{human}$ | 0.775 | 0.765 | 2/5 | -0.014 | -0.002 | — | 0.558 | 0.628 | 11/19 | 0.050 | 0.009 | 5.47E-01 | | | | | | |
| $A_{Crassvirales}$ | 0.816 | 0.789 | 3/5 | 0.027 | 0.011 | — | 0.561 | 0.650 | 12/19 | 0.053 | 0.013 | 1E+00 | | | | | | |
| $A_{SABC}$ | 0.810 | 0.799 | 4/5 | 0.021 | 0.020 | — | 0.540 | 0.633 | 12/19 | 0.032 | 0.004 | 1E+00 | | | | | | |
| **EV-D68** | | | | | | | | | | | | | | | | | | |
| $A_{total}$ | 0.470 | 0.415 | — | — | — | — | | | | | | | | | | | | |
| $A_{non\text{-}rRNA}$ | 0.486 | 0.489 | 2/5 | 0.016 | 0.000 | — | | | | | | | | | | | | |
| $A_{PMMoV}$ | 0.495 | 0.489 | 4/5 | 0.025 | 0.011 | — | | | | | | | | | | | | |
| $A_{ToBRFV}$ | 0.492 | 0.489 | 4/5 | 0.022 | 0.005 | — | | | | | | | | | | | | |
| $A_{Tobamovirus}$ | 0.489 | 0.476 | 3/5 | 0.020 | 0.011 | — | | | | | | | | | | | | |
| $A_{human}$ | 0.487 | 0.442 | 4/5 | 0.018 | 0.027 | — | | | | | | | | | | | | |
| $A_{Crassvirales}$ | 0.477 | 0.442 | 2/5 | 0.007 | 0.000 | — | | | | | | | | | | | | |
| $A_{SABC}$ | 0.466 | 0.418 | 1/5 | -0.003 | 0.000 | — | | | | | | | | | | | | |

**Supplementary Table S5. Per-pathogen wastewater PCR (WW-PCR) normalization summary.** For each PCR data source (WastewaterSCAN or National Wastewater Surveillance System (NWSS)), pathogen, and PCR normalization (rows), columns report per-site Spearman R between WW-PCR concentration and state-level Epic Cosmos clinical encounters or National Healthcare Safety Network (NHSN) Hospital Respiratory Data (HRD) admissions. The number of improved sites, and mean and median ΔR are reported relative to the raw-concentration baseline. P values are from a two-sided Wilcoxon signed-rank test on the paired per-site ΔR distribution (null: median ΔR = 0), Benjamini–Hochberg corrected across all (PCR source × pathogen × normalization × comparator) cells with a valid test; cells with fewer than six paired sites have a blank P value. Sites are restricted to the 5 WastewaterSCAN sites and 11 NWSS sites used in the main WW-MGS analysis (Supplementary Table S1; WastewaterSCAN takes precedence at sites with both). Because NHSN HRD reports influenza A and B as a single A+B-combined admissions series, WW-PCR Influenza A and Influenza B are both compared against the same NHSN flu admissions data. Empty cells indicate pathogen–comparator combinations with no data. PMMoV, pepper mild mottle virus.

| | **Epic Cosmos clinical encounters** | | | | | | **NHSN HRD hospital admissions** | | | | | |
|---|---|---|---|---|---|---|---|---|---|---|---|---|
| | Mean R | Med. R | Improved | Mean ΔR | Med. ΔR | p | Mean R | Med. R | Improved | Mean ΔR | Med. ΔR | p |
| **WastewaterSCAN** | | | | | | | | | | | | |
| **SARS-CoV-2** | | | | | | | | | | | | |
| Raw concentration | 0.728 | 0.684 | — | — | — | — | 0.538 | 0.595 | — | — | — | — |
| PMMoV-normalized | 0.762 | 0.749 | 4/5 | 0.034 | 0.037 | — | 0.496 | 0.547 | 2/5 | -0.042 | -0.049 | — |
| **Influenza A** | | | | | | | | | | | | |
| Raw concentration | 0.884 | 0.862 | — | — | — | — | 0.836 | 0.836 | — | — | — | — |
| PMMoV-normalized | 0.864 | 0.865 | 3/5 | -0.019 | 0.000 | — | 0.816 | 0.763 | 0/5 | -0.019 | -0.008 | — |
| **Influenza B** | | | | | | | | | | | | |
| Raw concentration | 0.732 | 0.725 | — | — | — | — | 0.651 | 0.652 | — | — | — | — |
| PMMoV-normalized | 0.651 | 0.630 | 1/5 | -0.081 | -0.050 | — | 0.538 | 0.593 | 1/5 | -0.113 | -0.115 | — |
| **RSV** | | | | | | | | | | | | |
| Raw concentration | 0.871 | 0.857 | — | — | — | — | 0.906 | 0.922 | — | — | — | — |
| PMMoV-normalized | 0.867 | 0.877 | 1/5 | -0.005 | -0.005 | — | 0.907 | 0.921 | 3/5 | 0.001 | 0.004 | — |
| **HMPV** | | | | | | | | | | | | |
| Raw concentration | 0.774 | 0.798 | — | — | — | — | | | | | | |
| PMMoV-normalized | 0.795 | 0.799 | 3/5 | 0.021 | 0.001 | — | | | | | | |
| **Norovirus** | | | | | | | | | | | | |
| Raw concentration | 0.650 | 0.682 | — | — | — | — | | | | | | |
| PMMoV-normalized | 0.589 | 0.567 | 0/5 | -0.061 | -0.059 | — | | | | | | |
| **Rotavirus** | | | | | | | | | | | | |
| Raw concentration | 0.601 | 0.601 | — | — | — | — | | | | | | |
| PMMoV-normalized | 0.437 | 0.437 | 0/1 | -0.164 | -0.164 | — | | | | | | |
| **National Wastewater Surveillance System** | | | | | | | | | | | | |
| **SARS-CoV-2** | | | | | | | | | | | | |
| Raw concentration | 0.744 | 0.777 | — | — | — | — | 0.722 | 0.749 | — | — | — | — |
| PMMoV-normalized | 0.755 | 0.776 | 1/4 | -0.092 | -0.071 | — | 0.835 | 0.838 | 4/4 | 0.227 | 0.197 | — |
| Flow-population-normalized | 0.681 | 0.687 | 0/11 | -0.063 | -0.044 | 1.95e-3 | 0.736 | 0.766 | 6/11 | 0.014 | 0.006 | — |
| **Influenza A** | | | | | | | | | | | | |
| Raw concentration | 0.826 | 0.805 | — | — | — | — | 0.822 | 0.825 | — | — | — | — |
| PMMoV-normalized | 0.697 | 0.683 | 0/4 | -0.129 | -0.127 | — | 0.754 | 0.750 | 0/4 | -0.068 | -0.079 | — |
| Flow-population-normalized | 0.763 | 0.740 | 0/4 | -0.063 | -0.065 | — | 0.790 | 0.790 | 1/4 | -0.032 | -0.024 | — |
| **RSV** | | | | | | | | | | | | |
| Raw concentration | 0.830 | 0.850 | — | — | — | — | 0.499 | 0.481 | — | — | — | — |
| PMMoV-normalized | 0.695 | 0.691 | 0/4 | -0.135 | -0.142 | — | 0.535 | 0.516 | 4/4 | 0.037 | 0.031 | — |
| Flow-population-normalized | 0.715 | 0.752 | 1/4 | -0.115 | -0.098 | — | 0.379 | 0.392 | 1/4 | -0.120 | -0.142 | — |
